# What approaches have been used to implement direct payments within health systems, and how do various factors influence the effectiveness of these approaches in supporting personalisation, governance, and equitable access to care: A Rapid Evidence Summary

**DOI:** 10.1101/2025.09.01.25334440

**Authors:** Elizabeth Gillen, Judit Csontos, Deborah Edwards, Adrian Edwards, Ruth Lewis

## Abstract

Continuing National Health Service Health Care (CHC) is a package of care for adults with significant primary health care needs who live in England or Wales. Currently, direct payments are not available for individuals receiving CHC in Wales. In contrast, in England, individuals in receipt of CHC can access direct payments as part of a broader system of Personal Health Budgets (PHBs), which offer choice and control over how their care is delivered. The *Health and Social Care (Wales) Act 2025* includes provisions enabling the introduction of direct payments for CHC in Wales, with implementation anticipated in 2026, subject to the development of supporting regulations and guidance

This review seeks to explore: what approaches have been used to implement direct payments within health systems, and how effective these approaches are in supporting personalisation, governance, and equitable access to care?

Searches were conducted on bibliographic databases from 2012 onwards to build upon previous work. Important pre-2012 grey literature evidence was also considered. The review included evidence published from 2010 to 2023.

The findings presented are based on the **8 review articles and 16 organisational reports**, some of which cover both health and social care.

**The literature lacks clear definitions** and consistent use of the terms related to direct payments and Personal Health Budgets (PHBs), often blurring the distinctions between different approaches. Where possible, findings have been drawn from the broader PHB literature, with relevant sections highlighted that directly address the implementation of direct payments.

Many of the **key elements for the successful implementation** of direct payments are similar across the different models of PHB implementation and include: Robust **support** and referral systems, **clear and accessible information** for recipients (patients and families), comprehensive **training and guidance** for staff involved in implementation to enhance knowledge and attitudes.

Policymakers should account for an initial adjustment period when assessing the impact of direct payments, as users and carers, as well as NHS staff, get used to any new arrangements and processes.

Researchers should carefully consider the timing of data collection in evaluations of direct payments, as early-stage data may disproportionately reflect implementation challenges rather than long-term outcomes. Longer-term follow-up (minimum of nine months) is essential to capture the full impact of personalised care, allowing users time to adjust, build confidence, and develop sustainable routines that reflect the intended benefits.

**Funding statement:** The authors and their Institutions were funded for this work by the Health and Care Research Wales Evidence Centre, itself funded by Health and Care Research Wales on behalf of Welsh Government.

## 1. CONTEXT / BACKGROUND

### 1.1 Background and purpose of the review

Personalised care is increasingly recognised as a cornerstone in the design and delivery of health and social care services in Wales. It reflects a strong policy commitment to ensuring that individuals, particularly those with complex or ongoing needs, have greater voice, choice, and control over how their care is arranged and delivered. One of the key mechanisms for enabling this personalisation in practice is the use of direct payments.

Direct payments involve providing individuals or their representatives with a monetary sum to arrange care in line with an agreed care and support plan. This approach empowers people to tailor their care to what matters most to them, offering greater flexibility, autonomy and control over how services are delivered. Support is often available to help individuals manage the associated responsibilities. Evidence suggests that direct payments can foster more responsive and personalised care that is better aligned with individual’s needs, routines, and preferences (Gadsby 2013). By enabling people to shape their care according to what matters most to them, direct payments promote and enable greater dignity, independence, and wellbeing.

In Wales direct payments have long been a central feature of personalised social care, administered by Local Authorities. However, they have not yet been extended into the healthcare system, which in Wales is overseen by Local Health Boards. This lack of continuity can result in a loss of control when individuals transition from social care into continuing healthcare disrupting established support arrangements and undermining person-centred practice (Welsh Government, 2025a; 2025b). This policy divergence has raised concerns among direct payments recipients, carers, and professionals about continuity and equity in service delivery.

To address these concerns, the Welsh Government has committed to introducing direct payments within Continuing NHS Healthcare. Following the passing of the Health and Social Care Act in Spring 2025, implementation of direct payments is anticipated in 2026. Unlike in England, where direct payments are one option within a broader personal health budget (PHB) model, Wales is taking a focused approach by introducing direct payments as a standalone mechanism to enhance personalisation in healthcare.

#### English approach

In England, Personal Health Budgets, which include as an option the receipt of a direct payment by those wishing to have a greater role in managing their own healthcare package, were piloted from 2009 and formally evaluated in 2012 through a mixed-methods study (Forder et al. 2012). The evaluation found that PHBs could be cost-effective, particularly for people receiving NHS Continuing Healthcare or mental health support, with improvements reported in social care-related quality of life (Forder et al. 2012). Subsequent research has reinforced the positive impact of PHBs on health, wellbeing, choice, and control, though achieving genuine personalisation requires significant shifts in NHS practice (Cooney et al. 2020; Ayoola & Butt, 2021). The NHS Mandate set a target of 50,000–100,000 PHB recipients by 2020/21, and by 2019/20, nearly 89,000 people had received one (NHS England, 2018; NHS Digital, 2020b). Initially restricted to specific groups, eligibility broadened under the NHS Long Term Plan to include individuals with learning disabilities, those under section 117 after-care, and users of bespoke support packages, with an ambition to reach 200,000 recipients by 2023/24 (NHS England, 2019a, 2022).

During the pilot, adults in receipt of NHS Continuing Healthcare (CHC) and living in their own homes were among those who benefited the most from Personal Health Budgets. Pilot sites enabled direct payments under the pilot arrangements, but the formal legal basis for making direct payments in CHC was only established later, through amendments to the NHS Act 2006 in 2014, which secured their availability beyond the pilot sites. Direct payments were also offered during the pilot to individuals with specific needs such as mental health conditions, long-term physical health problems, learning disabilities, or children and young people with complex needs (Irvine et al. 2011; Prabhakar et al. 2011; Jones et al. 2011; Forder et al. 2012; Davidson et al. 2012). In Wales, legislative change has now been made through the Health and Social Care (Wales) Act 2025, which amends the NHS (Wales) Act 2006 to enable the introduction of direct payments within NHS Continuing Healthcare.

Successfully implementing direct payments, whether as part of PHBs or as a standalone option, requires more than simply offering a cash alternative to traditional services. It demands thoughtful planning, supported by robust governance frameworks that ensure accountability, transparency and safeguarding. These frameworks must protect public funds, uphold quality of care and safeguard vulnerable individuals. To promote equity, it is essential to proactively address potential barriers that may affect uptake and implementation. Without appropriate support, individuals with limited capacity, social capital or digital literacy may struggle to navigate complex administrative systems. These considerations should be embedded into the design and planning stages to avoid exacerbating existing inequalities and to ensure that the benefits of direct payments are accessible to all eligible individuals.

A clear understanding is therefore needed of how direct payments have been implemented in other health systems and how effective these approaches are in supporting the core aims of personalisation, governance, and equitable access to care. This review seeks to explore:

*What approaches have been used to implement direct payments within health systems, and how effective are these approaches in supporting personalisation, governance, and equitable access to care?*

While our primary focus is on direct payments, we are also interested in relevant research on other forms of PHBs, including notional and third-party budgets. Including this broader evidence base will help us understand the differences in delivery models and identify transferable learning that may inform the effective implementation of direct payments in the Welsh context.

While this review draws extensively on evidence from the implementation of PHBs in England, it is important to recognise that the policy landscape in Wales is distinct. In Wales, PHBs are not part of the health system, and the focus is solely on the introduction of Direct Payments for Continuing NHS Healthcare. This distinction is crucial, as the legislative and operational frameworks governing direct payments in Wales will be unique and shaped by Welsh policy priorities. With the legislative framework now in place but infrastructure still to be developed, the Welsh approach to implementing direct payments in healthcare will need to be carefully designed to reflect local systems, priorities, and needs, while drawing on lessons from England. Therefore, while learning from PHB implementation in England is valuable, findings must be interpreted with this difference in mind.

### 1.2 Research question

To structure the review question and guide eligibility criteria, the SPICE framework was used. SPICE—standing for Setting, Perspective, Intervention, Comparison, and Evaluation (Booth 2006). The table below outlines the scope of the review. A more detailed summary of the methods used for conducting the Rapid Evidence Summary are provided in Section 5.

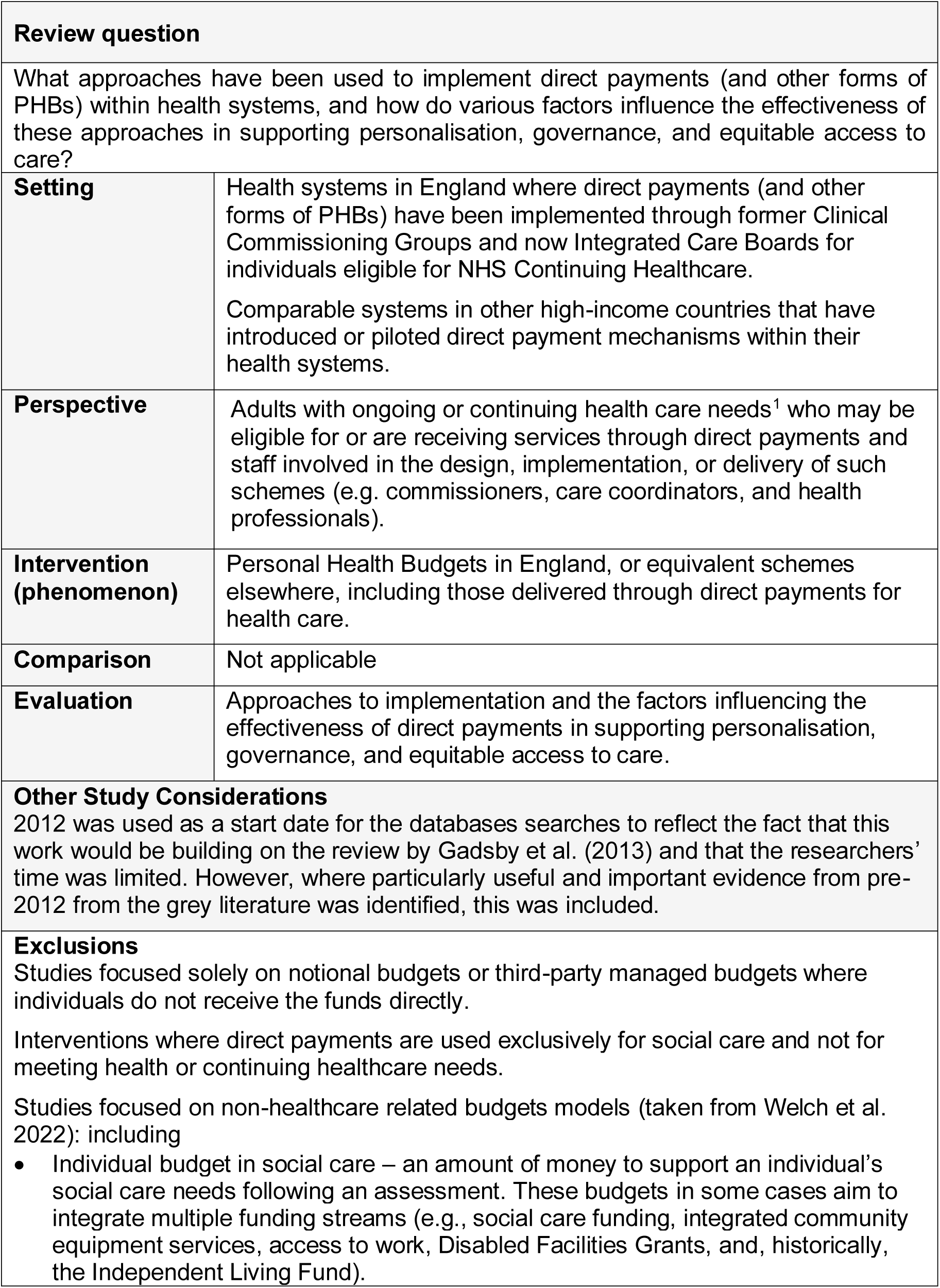
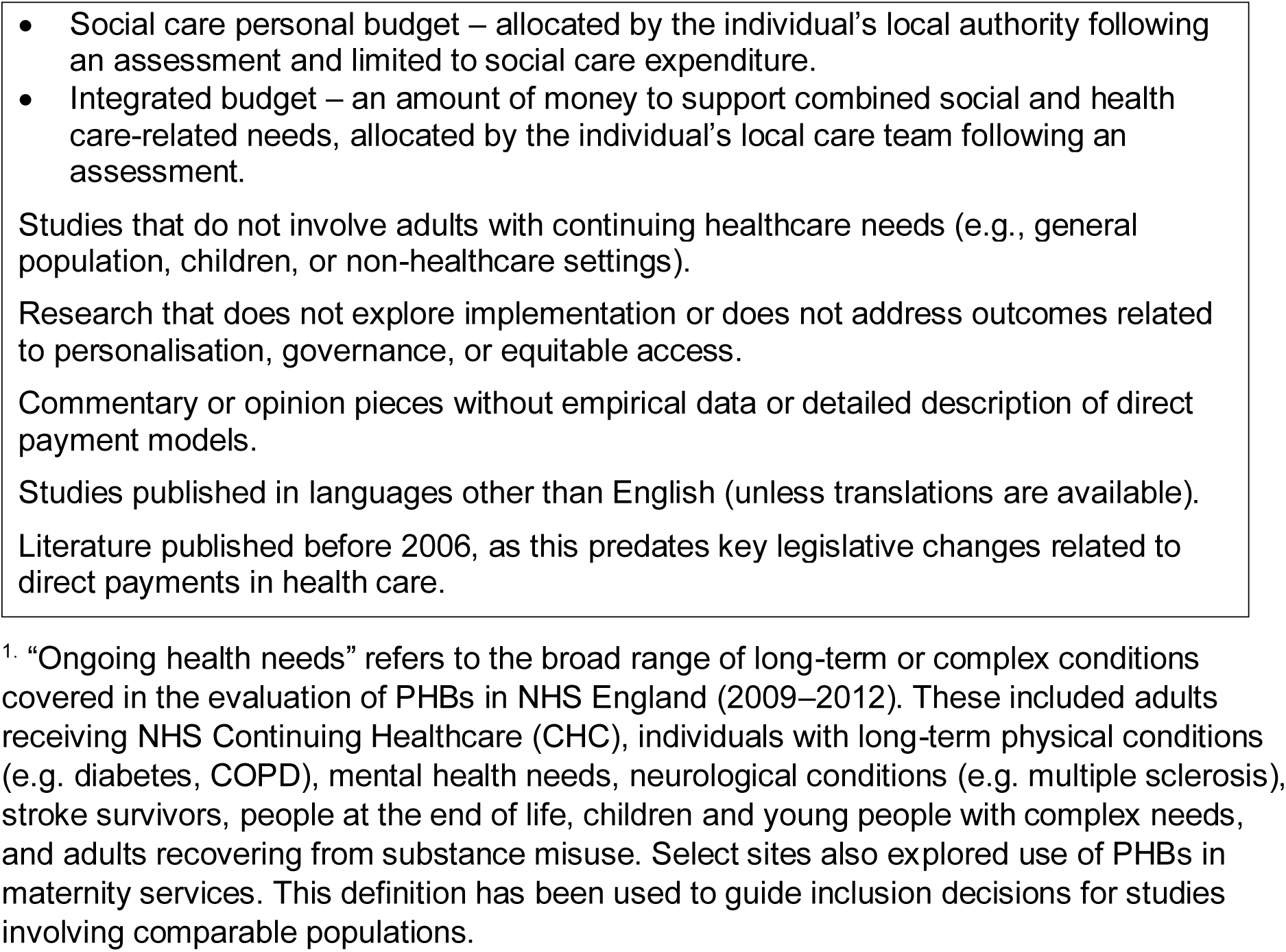

## 2. SUMMARY OF THE EVIDENCE BASE

### 2.1 Type and amount of evidence available

The type and amount of evidence retrieved is organised into systematic and rapid reviews, organisational reports and guidance documents and is summarised below. A summary of the findings of the included evidence is provided in Section 3.

#### 2.1.1 Systematic and rapid reviews

- We identified **two** rapid reviews (Health Foundation. 2010, Gadsby. 2013, Gadsby et al. 2013) and **six** systematic Review (Fleming et al. 2019, Lakhani et al. 2018, Micai et al. 2022, Robinson et al. 2022, Tompkins et al. 2018, Webber et al. 2014).
- **Six** of the reviews **examined both health and social care** (Fleming et al. 2019, Health Foundation. 2010, Gadsby. 2013, Gadsby et al. 2013, Lakhani et al. 2018, Micai et al. 2022, Robinson et al. 2022) while **two** focused **exclusively on healthcare** (Tompkins et al. 2018, Webber et al. 2014).
- There was **significant variation in terminology**, with terms such as ‘self-direction’, ‘individualised budgets’, ‘personal budgets’, ‘direct payments’, and ‘individualised funding’ often used interchangeably across different studies.
- The Health Foundation collated over 60 articles on **personal budgets across health and social care** in the UK and internationally. The objectives were to examine the international evidence on the impacts of personal health budgets on health outcomes, patient-centred care, and value for money; to explore whether personal health budgets are more effective for some groups of people; to identify where the majority of studies originate; and to review the UK evidence on individual budgets for social care. The review included studies published up to 2010 (Health Foundation 2010).
- A rapid review across 11 OECD countries that examined how **PHB models and self-directed support** were implemented, focusing on budget allocation, management, and governance. The review identified a range of models, from open to planned^2^, with some countries adopting hybrid approaches. The review included studies published up to 2012 (Gadsby 2013; Gadsby et al. 2013).
- A systematic review that synthesised evidence from Europe, the United States, Canada, and Australia to evaluate the effectiveness of **individualised funding** in improving **health and social care** outcomes for people with disabilities. It incorporates findings from 66 qualitative and three mixed-methods studies, examining stakeholder experiences with a particular emphasis on the challenges and facilitators encountered during the initial implementation phase of these interventions. The review covers studies published between 1992 and 2016 (Fleming et al. 2019).
- A systematic review that explored factors that influenced engagement with **self-directed models of health and social support** for people with various disabilities, including intellectual disabilities and degenerative diseases. Additionally, the review investigated how informed decisions were made and people chose services. The systematic review identified 18 reports, 15 primary studies and three review articles. Primary studies were conducted in six countries, six in the UK, five in the USA, and one each in Australia, Finland, New Zealand and Germany. Studies were published between 2012 and 2016 (Lakhani et al. 2018).
- A systematic review that focused on the **use of personal budgets** for people with mental health conditions or intellectual disability and included 29 studies published between 2013 and 2021 in four countries. The studies mainly originated from the UK and the USA, with 11 studies included from each country. Six studies were conducted in Italy and one study was included from Australia. Nineteen studies were qualitative by design exploring people’s, their carers and professionals’ experiences, whilst 10 studies had a quantitative design (Micai et al. 2022).
- A systematic review that explored the effects and costs of **personalised budgets** for people with **physical disabilities, intellectual and developmental disabilities, and mental health conditions.** The review focused on models where individuals had control over their care decisions, examining various forms of personalised budgeting. The review included a range of study designs, and the studies were conducted in high-income OECD countries, with 16 from the United States, four from England and three from Italy, covering the period from January 1985 to November 2022 (Robinson et al. 2022).
- A systematic review of four programmes across six studies focused on the implementation and delivery of **PHBs** initiatives for **drug and alcohol users** in England and the USA between 1990 and 2017. The review included both qualitative and mixed methods study designs (Tompkins et al. 2018).
- A systematic review that examined the effectiveness of **personal budgets for adults aged 18 to 65 with mental health problems**, including those with additional disabilities.

It included nine studies conducted in the United Kingdom and six in the United States, evaluating a range of models such as individual budgets, recovery budgets, personal budgets, **direct payments,** personal health budgets, and cash and counselling programmes. A variety of study designs were used to assess outcomes across different approaches to personalised budgeting (Webber et al. 2014).

#### 2.1.2 Organisational reports

- **Twelve** publications were identified that documented both the pilot phase and the subsequent national rollout of **PHBs within the NHS in England**. These included evaluations of the pilot (Davidson et al. 2012; Forder et al. 2012; Glendinning et al. 2013; Irvine et al. 2011; Jones et al. 2010a, 2010b, 2010c, 2011, 2013; Welch et al. 2013) as well as studies examining the later national implementation (Jones et al. 2017, 2018).
- The PHB pilot programme, launched by the Department of Health in 2009, was a national initiative in England aimed at testing the feasibility and impact of giving individuals greater control over their NHS-funded care.
  - A total of 64 sites participated in the programme, with 20 selected for in-depth evaluation and the remaining sites forming a broader comparison cohort (Jones et al. 2010a, b).
  - Personal health budgets were available for adults with continuing health care, long-term conditions, mental health needs, stroke survivors and parents of children with complex needs (Jones et al. 2010a, b).
- During the pilot, direct payments were not legally available for continuing health care recipients but could be offered to others, such as those with mental health needs, long-term conditions or learning disabilities under existing personalisation frameworks or temporary legal flexibilities.
- The evaluation sought to identify which implementation approaches were most effective in achieving positive outcomes for individuals (Jones et al. 2013; Forder et al. 2012).
- There were several focused studies conducted alongside the national evaluation of the PHB pilot programme. One strand examined the implementation process from the perspectives of organisational representatives (Jones et al. 2010b, 2010c) and budget holders (Irvine et al. 2010; Davidson et al. 2012). Other studies explored specific areas of interest:

- Jones et al. (2011) assessed the financial costs of planning and delivering PHBs across 20 pilot sites using different models;
- Welch et al. (2013) examined the use of PHBs in substance misuse services, focusing on outcomes such as impact, satisfaction, relapse, and implementation challenges; and
- Glendinning et al. (2013) investigated the application of PHBs in maternity care through in-depth case studies and interviews.
- The Department of Health commissioned a further study to explore the continued implementation of PHBs following the national pilot programme. Findings from this study are presented across two reports:

- The first report focuses on the perspectives of personal health budget leads, commissioners, and budget holders (Jones et al. 2017).
- The second report explores the views of service provider organisation managers and budget holders (Jones et al. 2018).
- A further **four** organisational reports contributed additional evidence on the implementation and outcomes of **PHBs in England and internationally** (Hatton and Waters 2015; Alakeson and Rumbold 2013; Skills for Care 2016, The Health Foundation 2011).
  - The Health Foundation (2011) examined the implementation of PHBs in the Netherlands through interviews with policymakers, carers, users, and experts. The case study was intended to inform UK policy development during England’s PHB pilot phase.
  - Hatton and Waters (2015) evaluated the experiences of personal health budget holders and family carers across 37 areas in England using the POET survey tool.
  - Skills for Care (2016) examined how PHB holders employing personal assistants **via direct payments** are supported across health and care systems.
  - The Nuffield Trust (Alakeson and Rumbold 2013) explored the implementation and implications of PHBs in England based on the data from the evaluation the national pilot programme (described above). The report was intended for commissioners and policy-makers in the UK health system to inform a wider roll-out of PHBs by highlighting practical, financial, and policy challenges.

#### 2.1.3 Guidance documents

We are highlighting **four** of the most recent guidance documents published by NHS England (relevant to the English context) that either focus specifically on direct payments for healthcare or, from the broader perspective of PHBs and **contain sections directly relevant to the implementation of direct payments**.

- A <u>guidance document</u> is intended to support Integrated Care Boards (ICBs) to understand and apply the direct payments for healthcare regulations (NHS England 2022a).
- A <u>guidance document</u> which is designed to help ICBs and other commissioners of health and care services understand the right to have a PHB for eligible groups. These include adults receiving NHS continuing healthcare, individuals receiving after-care under section 117 of the Mental Health Act (1983), people in receipt of NHS wheelchairs, and children and young people receiving continuing care (NHS England 2022b).
- A <u>guidance document</u> that clarifies the three deployment options for personal health budgets (direct payment, notional budget, and third-party budget) and outlines the responsibilities of both the individual and the commissioner. It also reinforces the requirement for ICBs to make all three options available (NHS England 2023a).
- A <u>guidance document</u> that aims to support practitioners and ICBs by outlining the decision-making process for delegating healthcare tasks from registered practitioners to personal assistants. It provides protocols to ensure delegation is safe and appropriate and clarifies roles, responsibilities, and lines of accountability (NHS England. 2023b).

## 3. KEY FINDINGS

This section draws on findings from both the eight review articles and the 16 organisational reports. Not all sources focused exclusively on healthcare, some addressed both health and social care. The existing review literature lacks consensus on the definitions and applications of key terms associated with direct payments and/or PHBs, often blurring the distinctions between different approaches. Where possible, findings have been drawn from the broader PHB literature, with relevant sections highlighted that **directly address the implementation of direct payments.** Many of the challenges and facilitators identified are similar across the different models of PHB implementation. Key findings are highlighted below, with the full results of relevance presented within Tables 2 to 5 (presented in Section 6).

### 3.1 Personalisation

This section explores the role of personalised delivery in enhancing choice, control and flexibility through direct payments and/or PHBs and examines how these elements relate to outcomes such as quality of life. It also considers some of the practical challenges that can arise in implementing personalised approaches.

#### 3.1.1 Choice and control

- International evidence shows that well implemented personal budgets for **health and social care** can enhance choice, control, and confidence in managing care (Health Foundation 2010).
- All interventions aimed to enhance user choice and control, though the level of personalisation varied across programmes (Robinson et al. 2022).
- Personal budgets increased choice; individuals appreciated having a wider range of services and more discretion over how time and resources were used (Webber et al. 2014).
- Participants felt more in control of their lives and support, with increased confidence and empowerment (Webber et al. 2014).
- **Direct payments** were widely seen as the most empowering option, with some budget holders and frontline staff viewing them as the only true route to full control (Jones et al. 2010c).
- Some participants viewed **direct payments** as enhancing personalisation and giving individuals greater control over their care (Welch et al. 2013).
- Users valued the control **direct payments** offered (Jones et al. 2017).
- Budget holders experienced greater control over their care arrangements (Jones et al. 2018).
- Funding models that directly allocated budgets to users enabled access to services and activities that might otherwise have been out of reach (Lakhani et al. 2018).
- Self-direction can increase choice and control for disabled people, helping them align services with their needs and aspirations (Lakhani et al. 2018).
- Use of personal budgets by people with mental health conditions has been associated with increased choice and control (Micai et al. 2022).

#### 3.1.2 Outcomes of personalisation

- Personalised budgets were generally associated with improved quality of life and care satisfaction for both service users and carers (Robinson et al. 2022).
- International evidence shows that well implemented personal budgets are associated with improved quality of life and satisfaction with care (Health Foundation 2010).
- Reported benefits included improved quality of life, physical health, mental health, and relationships, though not consistently across all participants (Webber et al. 2014).
- Delivery of PHBs through **di**r**ect payments** had a positive impact on carers’ well-being (Jones et al. 2010c).
- PHBs were seen to improve quality of life through greater choice, control, and tailoring of services to personal needs and circumstances (Jones et al. 2010c).
- Users reported improved health outcomes from using **direct payments** (Jones et al. 2017).
- Users reported improved independence, dignity, and overall quality of life (Jones et al. 2018)
- Improved responsibility and awareness, quality of life, independent living, employment, and clinical, psychological, social, and daily outcomes have been observed among patients with mental health conditions who utilise personal health budgets. These positive outcomes are associated with increased choice and control, patient empowerment, timely and appropriate access to treatment, active involvement of carers, and collaborative care planning with professionals and stakeholders (Micai et al. 2022).
- **Direct payments** managed by family or friends were more often linked to positive impacts on living arrangements and feeling supported with dignity, compared to those managed by the individuals themselves (Hatton and Waters 2015)
- Some negative effects were reported, including increased cognitive burden among individuals with mental health or cognitive conditions, highlighting the need for more tailored approaches (Robinson et al. 2022).
- Evidence does not clearly support the assumption that greater choice automatically leads to better outcomes or lower costs for PHB recipients (Gadsby 2013; Gadsby et al. 2013).

#### 3.1.3 Flexibility and responsiveness

- International evidence also links personal budgets in **health and social care** to more flexible and responsive services overall (Health Foundation 2010).
- Personal budgets for individuals with mental health problems increased the range of service options and allowed for more flexible use of time and resources (Webber et al. 2014).
- **Direct payments** offered more respite and flexibility, but carers were also concerned about increased responsibility (Jones et al. 2010c).
- **Direct payments** were seen as a way to increase flexibility and improve responsiveness, especially for smaller or ad-hoc purchases (Welch et al. 2013).
- Budget holders experienced greater flexibility over their care arrangements (Jones et al. 2018).
- Flexibility contributed to improved service quality and reduced staff turnover for some users (Lakhani et al. 2018)
- Flexibility was a key benefit, offering greater choice, control and responsiveness to individual needs both in the type and timing of support, and in how funding was used (Flemming et al. 2019).
- Staff delivering personal budgets to people with drug and alcohol problems reported gaining a better understanding of their needs, leading to more trusting relationships, greater flexibility, and increased job satisfaction (Tompkins et al. 2018).

#### 3.1.4 Challenges in personalised delivery

- Evidence on the extent of meaningful choice in self-directed programmes was mixed due to variation in implementation, support, information, and funding controls (Lakhani et al. 2018).
- Reduced service quality was noted as a potential drawback of personal budgets, particularly where unqualified carers were employed in unregulated environments (Lakhani et al. 2018).
- Some individuals, especially those who struggled to express their needs, found directing their own care challenging. Mental health service users often felt less in control than other social care groups despite receiving a personal budget (Webber et al. 2014).
- The assumption that increased choice automatically results in greater autonomy is overly simplistic and not well supported by evidence from recipients of PHBs (Gadsby 2013; Gadsby et al. 2013).
- People with drug and alcohol problems had varied experiences of personalisation, with many unaware of their PHB’s value and often having limited control, as budgets were frequently managed notionally or by keyworkers (Tompkins et al. 2018).

### 3.2 Use of personal health budgets

This section describes the **motivations** for choosing direct payments and/or personal health budgets, followed by a description of **how these are used in practice**, including the types of purchases made and the management arrangements involved.

#### 3.2.1 Motivations for choosing direct payments

- Motivations for choosing **direct payments** included lifestyle fit, greater choice and flexibility, avoiding third-party fees, prior experience, or enjoyment of administrative tasks (Irvine et al. 2011).
- Most people using **direct payments** felt it was the right option for them (Davidson et al. 2012).

#### 3.2.2 Types of purchases and management arrangements

- **Direct payments** enabled personalised care planning, including access to non-traditional services such as complementary therapies (Jones et al. 2018).
- Budgets enabled access to extra-curricular activities aligned with users’ interests and aspirations (Lakhani et al. 2018).
- Most people managed their PHB through **direct payments** to themselves or via a family member or friend, while fewer used service providers, brokers, or NHS/council-managed options (Hatton and Waters 2015).
- Some participants reported that the **health and social care** programmes approved by funding agencies were limited in scope (Lakhani et al. 2018).
- Personal health budgets for people with drug and alcohol problems were used not only for core treatment but also for broader lifestyle, educational, and psychosocial supports that individuals felt contributed to their recovery (Tompkins et al. 2018).

### 3.3 Equitable access

This section examines the extent to which **equitable access** to direct payments and/or PHBs is achieved across different population groups. It includes findings on differential access and benefit, selection bias, barriers to timely and informed access, and challenges in delivering personalised support across diverse populations.

#### 3.3.1 Differential access and benefit

- Self-directed approaches appeared more beneficial for those with family support, middle-class backgrounds, or higher educational resources (Lakhani et al. 2018).
- Some people may benefit more from personal budgets across **health and social care,** especially those with good support, advice, or higher confidence and skills (Health Foundation 2010).
- Concerns about equity in PHB programmes stem from the idea that those with higher education and stronger social networks are better equipped to benefit, while others may struggle. Limited research means the impact on health inequalities remains unclear (Gadsby 2013; Gadsby et al. 2013).
- **Direct payments** were seen as having the potential to improve equity for Black and minority ethnic budget holders by enabling access to more culturally and linguistically appropriate support, including family-led services (Jones et al. 2010c).

#### 3.3.2 Selection bias and underrepresentation

- Evidence from England suggests health professionals may favour younger, more educated individuals for PHBs, with underrepresentation of older adults and minority ethnic groups in the PHB pilot, pointing to potential selection bias in implementation (Gadsby 2013; Gadsby et al. 2013).

#### 3.3.3 Barriers to timely and informed access

- Delays, unclear protocols, and lack of guidance limited timely and fair access to PHBs for people using drug and alcohol services. Service users reported uncertainty about eligible uses and wanted clearer information during care planning (Tompkins et al. 2018).
- Decision-making was affected by access to information, support, location, and socioeconomic factors (Lakhani et al. 2018).
- People in rural or remote areas faced greater barriers—limited services, higher costs (e.g. travel), and difficulties managing care (Lakhani et al. 2018).
- When users were restricted to a list of government-approved providers, informed choice was undermined (Lakhani et al. 2018).

#### 3.3.4 Challenges in personalised delivery

- Staff reported logistical challenges in delivering individualised support, particularly in meeting diverse expectations and accommodating socio-demographic differences (Fleming et al. 2019).

### 3.4 Implementation challenges

This section describes key implementation challenges associated with direct payments and wider PHBs, including system and cultural change, process complexity, uptake and cost management, and workforce impact.

#### 3.4.1 Structural and cultural change

- Implementation takes time and often requires cultural and structural change within existing systems (Gadsby 2013; Gadsby et al. 2013).
- Successful engagement among service users with self-directed programmes requires a cultural shift across service providers (Lakhani et al. 2018).
- Traditional service models and provider attitudes can resist the shift toward personalised approaches (Gadsby 2013; Gadsby et al. 2013).
- Aligning personal budgets with current processes can be complex (Gadsby 2013; Gadsby et al. 2013).
- The systems in place were often cumbersome and duplicated work. They tended to prioritise targets and cost-efficiency over the actual support provided, creating barriers to personalised care (Flemming et al. 2019).
- The absence of national systems for resource allocation placed a burden on families to negotiate access to funding (Fleming et al. 2019).
- Variation across local areas in what personal health budgets could fund led to confusion and perceived inequities in delivery (Jones et al. 2018).
- Limited awareness of personal health budgets among patients and providers was a barrier to wider service development and uptake (Jones et al. 2018).
- Service users and their families should be central, seen as capable of making care choices, and supported with accessible resources (Lakhani et al. 2018).
- Commissioners must plan for decommissioning services not chosen by PHB holders to prevent duplication and market shrinkage. At the same time, they need to support the growth and diversification of the provider market using strategies that avoid destabilising existing services (Alakeson and Rumbold 2013).
- Infrastructure is needed for budget setting, care planning, and monitoring; using existing systems may support efficiency (Alakeson and Rumbold 2013).
- Integrating PHBs with social care budgets through a coordinated ‘dual carriageway’ model may support service integration without structural merger (Alakeson and Rumbold 2013).
- Risk of a ‘postcode lottery’ highlights the need for consistent national implementation (Alakeson and Rumbold 2013).

#### 3.4.2 Uptake and resource management

- Individual take-up is often slower than expected (Gadsby 2013; Gadsby et al. 2013).
- Costs must be carefully managed, primarily through limits on individual budget allocations, alongside the application of eligibility criteria (Gadsby 2013; Gadsby et al. 2013).
- Policymakers need to plan for the set-up and transition costs of individualised funding models, which are often underestimated (Fleming et al. 2019).
- In many systems, individuals are expected to contribute or cover funding shortfalls (Gadsby 2013; Gadsby et al. 2013).
- Participants experienced financial hardship due to hidden costs associated with managing personal budgets (Fleming et al. 2019).
- All interventions involved a transitionary period and a major challenge during implementation was the lack of national systems for resource allocation which placed a burden on families to negotiate access to funding (Fleming et al. 2019).

#### 3.4.3 Workforce and delivery pressure

- Viewing service users as consumers may motivate providers to improve service standards (Lakhani et al. 2018).
- Some professionals viewed the management of personal budgets as a challenging yet important aspect of their role (Micai et al. 2022).
- It was recognised that there could often be blurring between the positive and the negative, where one person could feel empowered by directly employing support whilst another may find this stressful (Flemming et al. 2019).
- Staff noted that delivering PHBs to people with drug and alcohol problems was time-consuming and often involved extra responsibilities beyond their usual role (Tompkins et al. 2018).

### 3.5 Factors that enable or hinder successful implementation

This section describes key enablers of successful implementation of direct payments and/or PHBs, including brokerage and support mechanisms, accessible information, effective training and organisational development. It also considers potential challenges such as bureaucracy and administrative burden, recruitment and staffing and communication barriers.

#### 3.5.1 Brokerage and support mechanisms

- Brokerage, which can involve advice, information, and hands-on help, enables individuals to manage their care more effectively (Gadsby 2013; Gadsby et al. 2013).
- Brokerage and signposting support are important and may be most effective when delivered by the voluntary sector or independently of the services being offered (Health Foundation 2010).
- Effective support or signposting mechanisms are key to enabling individuals to understand and manage **direct payments** (Health Foundation 2010).
- independent support services for budget holders are highly valued and are linked to more positive, person-centred outcomes (Gadsby 2013; Gadsby et al. 2013).
- Paid supporters are needed who have strong communication and facilitation skills to help individuals identify their short- and long-term goals and understand the steps needed to achieve them, especially when doing so for the first time (Fleming et al. 2019).
- Strong trusting and collaborative relationships (paid and unpaid) that facilitate information sourcing, staff recruitment, network building and administrative and agency support (Fleming et al. 2019).
- Budget holders greatly valued the support provided from PHB lead officers or support workers including help with advertising, shortlisting, and conducting interviews (Davidson et al. 2012).
- Budget holders for **direct payments** either managed employment^3^ tasks such as tax and National Insurance themselves or opted to use an agency. Those who chose to manage the tasks independently sometimes felt daunted and drew on the support from professionals, friends or family (Davidson et al. 2012).
- It is recommended that procurement and recruitment support be provided for personal health budget holders, particularly those using **direct payments**, as this support is valued by recipients (Forder et al. 2012).
- For those new to managing **direct payments,** ongoing professional support was considered essential. (Jones et al. 2017).
- Independent third-party advisors, such as support brokers and social workers, played a valuable role in supporting individuals to make informed decisions (Lakhani et al. 2018).
- Seeking advice from trusted individuals, such as parents or peers in similar circumstances, was identified as a key factor in supporting choice. However, Internal family conflicts could hinder decision-making, highlighting the need for clear roles and authority within **self-directed support** arrangements. (Lakhani et al. 2018).
- Individuals who received **direct payments** themselves tended to plan more independently, while those whose payments were managed by family, friends, or brokers were more likely to rely on support from those individuals (Hatton and Waters 2015).
- People receiving payments directly found it easier to manage and access support, while those with broker-managed budgets found it easier to make changes to their support (Hatton and Waters 2015).
- A strong network of support, including family, friends, colleagues, and paid coordinators or brokers, was key to sourcing information, recruiting staff, expanding connections, and managing administrative tasks (Fleming et al. 2019).

#### 3.5.2 Employment of personal assistants and carers

- Employed carers or personal assistants were often friends, relatives, or former care staff. (Davidson et al. 2012).
- Holding interviews for carers and personal assistants in professional settings rather than at home was described as a significant relief (Davidson et al. 2012).
- Challenges included recruiting suitable carers and personal assistants locally, covering advertising costs, and finding applicants willing to work on payroll rather than cash-in-hand (Davidson et al. 2012).
- Breakdowns in existing care arrangements of carers and personal assistants and staffing difficulties highlighted the need for accessible back-up and recruitment support (Davidson et al. 2012).
- Budget holders varied in their views on Criminal Records Bureau (CRB – now known as Disclosures and Barring Service - DBS) checks; some felt they were unnecessary when employing relatives or pre-checked staff, while others valued support from professionals in arranging them (Davidson et al. 2012).
- NHS organisations, local authorities, and other CRB holders should work together to address recruitment and retention challenges for personal assistants (Skills for Care 2016).
- National-level work is needed to understand how new models of personal assistant employment may affect current care systems (Skills for Care 2016).
- Core training should be provided to personal assistants, linked where appropriate to the Care Certificate standards (Skills for Care 2016).
- Healthcare systems should plan for increased demand by creating roles to oversee personal assistant training, sign-off, and review of competence (Skills for Care 2016).
- Personal assistants should have access to peer support or, where not feasible, a neutral point of contact for workplace or human resources issues, separate from employer support (Skills for Care 2016).

#### 3.5.3 Information and communication for recipients of direct payments and/or personalised health budgets

- Accessible information for potential budget holders is a key factor in the successful implementation (Health Foundation 2010).
- Recipients of PHBs often encounter unfamiliar systems and responsibilities, making timely access to support and information essential. (Gadsby et al. 2013).
- Positive experiences were associated with support that was knowledgeable, accessible, and responsive to individual needs (Gadsby et al. 2013).
- Effective implementation requires accessible information and tailored assistance, including practical help with managing employment, contracts, and finances (Gadsby et al. 2013).
- Details of **direct payment** options were not always clearly explained, including whether full annual budgets would be paid upfront (Irvine et al. 2011).
- Clear information is needed both during and after care planning, as questions may arise once PHBs are in use (Irvine et al. 2011).
- Limited access to accurate, accessible information and advice is a persistent barrier to informed decision-making (Lakhani et al. 2018).
- Inadequate or misleading information can lead to poor choices and the inefficient use of resources (Lakhani et al. 2018).
- Common concerns included inaccurate, mixed, or inaccessible information, highlighting the need for clearer guidance on available supports, how to access them, and what funding could be used for (Fleming et al. 2019).

#### 3.5.4 Training and support for healthcare staff involved in Implementing direct payments and/or personal health budgets?

- Training and guidance to improve the knowledge and attitudes of frontline healthcare staff and local authority leadership is important (Health Foundation 2010).
- Training for healthcare staff should focus on managing change, improving assessments and promoting equality and challenging assumptions about who is suitable for personal budgets (Gadsby 2013; Gadsby et al. 2013).
- Timely training and education for frontline healthcare are essential to build the knowledge and confidence required to implement **direct payments** effectively (Fleming et al. 2019).
- Healthcare staff felt underprepared to support **direct payments** due to lack of training and guidance (Jones et al. 2010c).
- Clinical commissioning groups (now Integrated Care Boards) should develop local frameworks to support the delegation of healthcare tasks to personal assistants including training and competence assessment (Skills for Care 2016).
- Significant concerns were raised regarding the limited capacity of small local organisations to meet increasing demand with no alternative structures in place. (Flemming et al. 2019).
- Greater investment in education and training is needed to support stakeholder buy-in and understanding of individualised funding and its implementation (Fleming et al. 2019).

#### 3.5.5 Bureaucracy and administrative burden

- Participants often found managing **direct payments** stressful, particularly during early implementation, due to the complexity, bureaucracy, and significant legal and administrative responsibilities involved. This was especially challenging for those without prior managerial experience, who had previously held more passive roles in traditional services (Jones et al. 2017, Jones et al. 2018; Flemming et al. 2019; Micae et al. 2022).
- While individuals with organisational skills, assertiveness, and relative experience found the process more manageable, reduced professional support since the pilot programme left many users less equipped to handle these responsibilities effectively (Jones et al. 2017, Jones et al. 2018).
- Initial issues included setting up bank accounts, managing payments, and delays in funds being deposited or notified (Davidson et al. 2012).
- To help manage the demands of employment administration, some individuals chose to use payroll services, which helped reduce stress and streamline the process (Irvine et al. 2011).

### 3.6 Eligibility

This section outlines how **eligibility for direct payments and/or PHBs** is determined within health systems. It summarises formal criteria, local variation in assessment practices, staff perspectives and concerns, and considerations specific to safeguarding and the inclusion of particular groups such as people with substance misuse issues.

#### 3.6.1 Formal eligibility criteria and variation in practice

- Eligibility for **direct payments across health and social care** is limited to individuals assessed as needing community care services and who are considered willing and able to manage the payments, with support if needed (Health Foundation 2010).
- However, in practice, local authority teams have sometimes applied eligibility across **health and social care** selectively, with staff perceptions influencing who is encouraged to take up **direct payments**, often favouring younger disabled people (Health Foundation 2010).
- Internationally, the ways people accessed funding varied and was influenced by local policy priorities and infrastructure (Fleming et al. 2019).

#### 3.6.2 Inconsistencies in assessment and budget setting

- Care planning and budget setting for personal health budgets in the UK varied widely across **England’s PHB programme** pilot sites. Limited cost data made budget setting difficult, leading to diverse approaches with no clearly superior method (Gadsby 2013; Gadsby et al. 2013).
- Healthcare staff were unsure who to target for PHBs, with inconsistent guidance and differing views on service user readiness and capacity (Tompkins et al. 2018).
- Eligibility assessments often focused not only on the individual’s needs but also on the strength of their support network, which can introduce bias and limit access (Fleming et al. 2019).

#### 3.6.3 Staff uncertainty and ethical concerns

- Staff were uncertain about which people with drug and alcohol problems should be offered personal budgets and at what stage in their treatment (Tompkins et al. 2018).
- Staff delivering personal budgets to people with drug and alcohol problems reported moral and ethical concerns about whether this population could make appropriate spending decisions (Tompkins et al. 2018).

#### 3.6.4 Eligibility safeguards and discrimination

- Concerns were raised about offering **direct payments** to people with substance misuse issues, citing risks such as relapse, exploitation, and financial mismanagement (Welsh et al. 2013).
- Suggested safeguards when working with people with substance misuse issues included using care navigators or trusted third parties to manage funds or requiring abstinence before eligibility (Welsh et al. 2013).
- Requiring abstinence or third-party management of budgets was viewed by participants working with people with substance misuse issues as potentially discriminatory (Welsh et al. 2013).
- Those supporting individuals with substance misuse issues argue that, with appropriate safeguards, **direct payments** can promote autonomy and personal responsibility (Welsh et al. 2013).

### 3.7 Governance

This section outlines a range of accountability and risk management factors that need to be considered to successfully implement direct payments and points to some structural responses and lessons.

#### 3.7.1 Accountability and risk management

- Concerns about accountability and risk management for **personal budgets across health and social care** were raised, particularly when individuals employ their own staff or use unregistered providers (Health Foundation 2010).
- Safeguarding risks were noted in relation to informal care and the employment of family members or unqualified carers through **direct payments** (Gadsby 2013; Jones et al. 2010c; Lakhani et al. 2018).
- Several implementation challenges were highlighted, including the complexity of budget management potentially increasing exposure to risk, concerns that care organisation-led approaches may prioritise cost-efficiency over user choice, and reports of reduced service quality in settings where unqualified carers operated without formal regulation (Lakhani et al. 2018).
- Staff were concerned about the potential for the misuse of funds (Flemming et al. 2019; Jones et al. 2010c).
- In response to issues with **intermediary agencies** in the Dutch PHB system, including fraud and aggressive marketing, the government introduced reforms such as banning **direct payments** to intermediaries and implementing a voluntary code of practice (Alakeson and Rumbold 2013).

#### 3.7.2 Clarity of roles and decision-making

- Governance of PHBs varied widely across sites, with unclear roles and decision-making authority complicating efforts to balance user flexibility with strong oversight and accountability (Gadsby 2013).
- There was a lack of clarity around who approved PHB requests and how decisions were monitored for people with drug and alcohol problems, leading to delays and inconsistencies. Staff highlighted the need for clearer governance structures to support this population (Tompkins et al. 2018).
- There was confusion about appeals processes and who was accountable if things went wrong (Jones et al. 2010c).

#### 3.7.3 Formal governance structures

- Sites with formal governance structures (e.g. boards or steering groups) achieved more coherent implementation by clarifying roles, ensuring consistency, and supporting strategic oversight (Gadsby 2013).

### 3.8 Areas of uncertainty

- Existing literature lacks consensus on the definitions and applications of key terms associated with PHBs, often blurring the distinction between the different approaches, limiting clarity in interpreting the specific impact of direct payments.
- Despite national guidance, the design and delivery of PHBs can vary widely due to differing local interpretations of roles, responsibilities, and spending rules. This flexibility, while empowering, has led to inconsistent practices across regions, highlighting the need for clearer frameworks to ensure equitable and effective personalised care.
- When evaluating the impact of PHBs and direct payments, it’s important for policymakers to consider the initial adjustment period experiences by many users and carers. To capture more accurate outcomes, studies should include follow-ups of at least nine months and, ideally, collect data at multiple time points over an extended period.
- Due to time constraints, this summary does not fully reflect the experiences or outcomes for carers, which remain an important but underexplored aspect of PHB evaluations.

## Data Availability

All data produced in the present study are available upon reasonable request to the authors

## Abbreviations

COPD: Chronic obstructive pulmonary disease
CRB: riminal records bureau
ICB: Integrated care boards
NHS: National Health Service
PA: Personal assistants
PHB: Personal health budgets

## Glossary

### Continuing healthcare

“Continuing Health Care is a complete package of ongoing care arranged and funded solely by the NHS through local health boards, where an individual’s primary need has been assessed as health-based. Continuing Health Care is one element of a range of services that local authorities and NHS bodies need to have in place to support people with health and social care needs. Continuing Health Care is one aspect of care which people with complex needs may need as the result of disability, accident or illness to address both physical and mental health needs.”. (Welsh Government 2022, p.4.)

### Direct payments

Direct payments for healthcare are monetary payments in lieu of services. The money is individual (or their representative) receives the money directly and takes full responsibility for purchasing and managing services in line with their agreed care plan. Direct payments give individuals greater choice and control over their care, but they do not remove or alter the NHS’s legal duty of care to every person receiving services. (NHS England, 2022b).

### Notional budget

Where the NHS retains control of the funds but works with the individual to plan and arrange care based on their choices. This option offers personalisation without the need to handle the money directly (NHS England, 2022b).

### Personal assistants

Direct payments for healthcare can be used to pay for a personal assistant to carry out certain healthcare tasks. PAs can only carry out delegated healthcare tasks (e.g. clinical interventions) if these are included in the care plan and they receive proper training, supervision, and competency assessment (Skills for Care, 2023).

### Personal health budgets

Personal health budgets as implemented in England, use NHS funding to create an individually agreed personalised care and support plan that offers people of all ages greater choice and flexibility over how their assessed health and wellbeing needs are met (NHS England, 2022b). A PHB can be managed in three ways, via direct payments, a third-party budget, or a notional budget, or through a combination of these.

### Third-party budget

Where an independent organisation manages the funds on behalf of the person and arranges care according to their preferences, this allows for choice and flexibility without the administrative burden (NHS England, 2022b).

## 5. RAPID EVIDENCE SUMMARY METHODS

A list of the resources searched during this Rapid Evidence Summary is provided within Appendix. Searches were limited to English-language publications and did not include searches for primary studies as secondary research relevant to the question was found. Searches were limited from 2012 to current date (June 2025) following a review already published in this area by Gadsby et al. 2013. However, where particularly useful and important evidence from pre-2012 from the grey literature was identified, this was not excluded. Search hits were screened for relevance by a single reviewer.

Priority was given to robust evidence synthesis using minimum standards (systematic search, study selection and appropriate synthesis). Quality appraisal was not undertaken as part of this Rapid Evidence Summary and consequently the included research may vary in quality. Citation, recency, evidence type, document status and key findings were tabulated for all relevant secondary research identified in this process.

Our Rapid Evidence Summaries are generally based on key information extracted from the abstracts of included material. However, the nature of the current review question and included material meant that the relevant key information was rarely reported in the abstract. Rather this was frequently embedded in lengthy and complex organisational reports and systematic reviews. Therefore, in light of time constraints of this evidence summary, a single reviewer used artificial intelligence tools Microsoft Co-Pilot or ChatGPT 4.0 to extract the relevant key information included in these documents and then to assist in concisely summarising their content. All extracted and summarised data was subsequently checked for accuracy to ensure clarity, consistency, and reliability.

|  |  |
| --- | --- |
| **Date of Search** | June 2025 |
| **Search Concepts Used** | direct payment* OR personal health budget* OR personalisation OR personalization OR individual health budget*<br>(For full search audits see Appendix 1) |
| **Search Completed by** | Elizabeth Gillen 10.06.2025 |

## 6. EVIDENCE

**Table 1:**
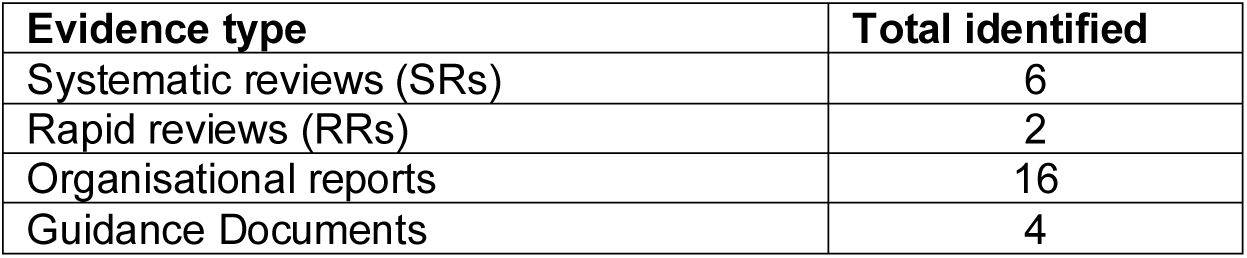
Summary of review evidence identified.

| **Evidence type** | **Total identified** |
| --- | --- |
| Systematic reviews (SRs) | 6 |
| Rapid reviews (RRs) | 2 |
| Organisational reports | 16 |
| Guidance Documents | 4 |

A more detailed summary of included evidence can be found in Tables 2 to 5.

**Table 2:**
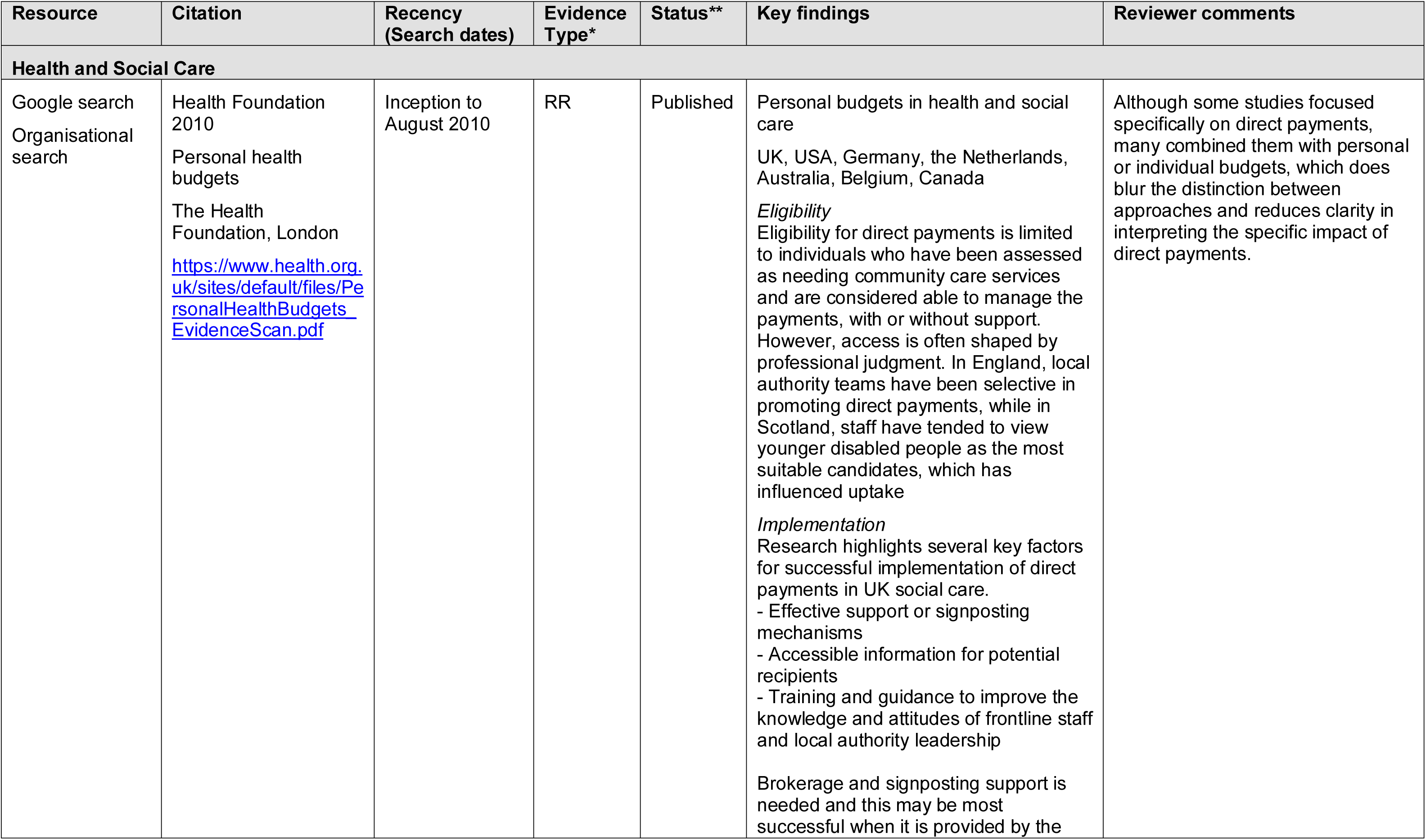
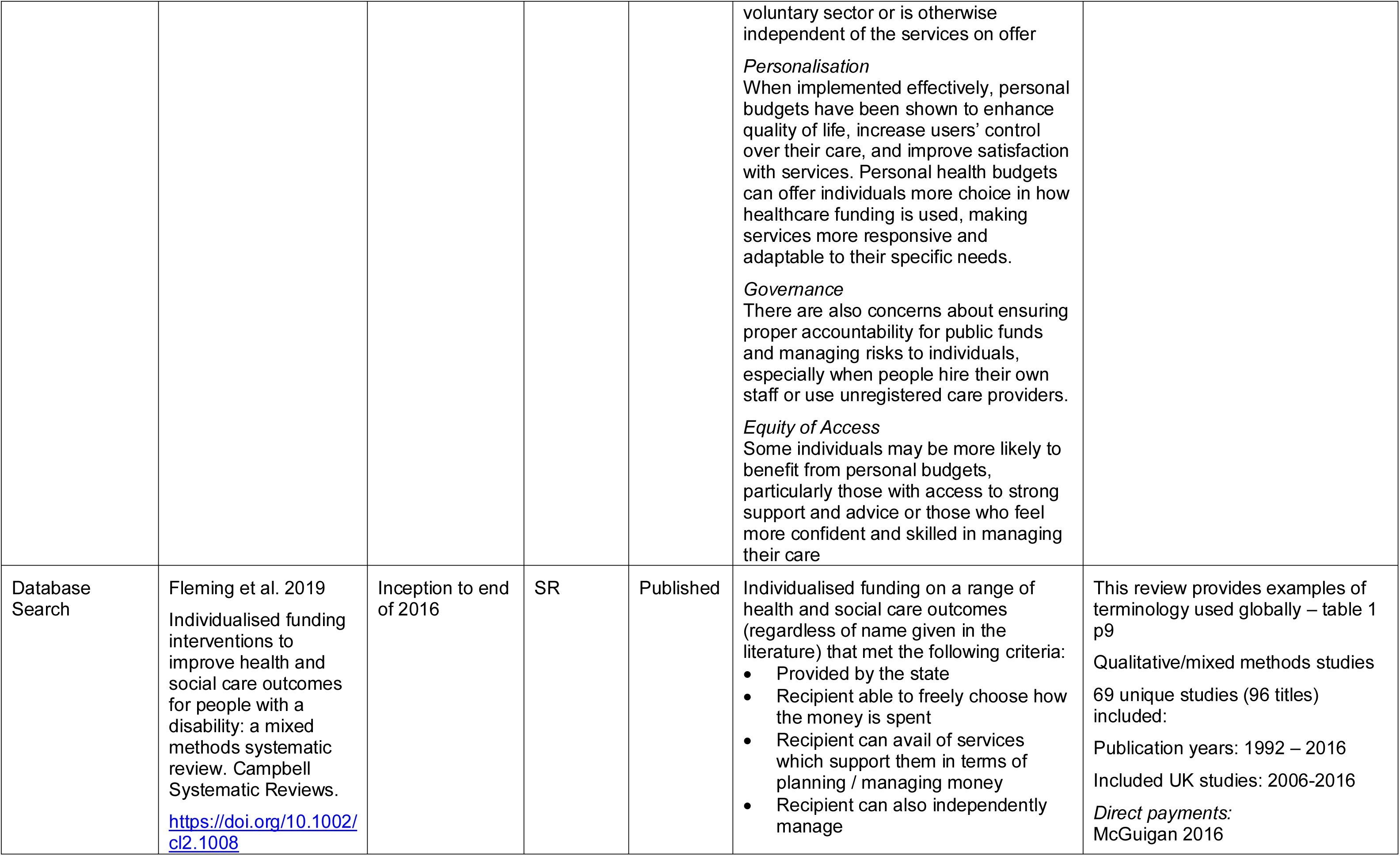
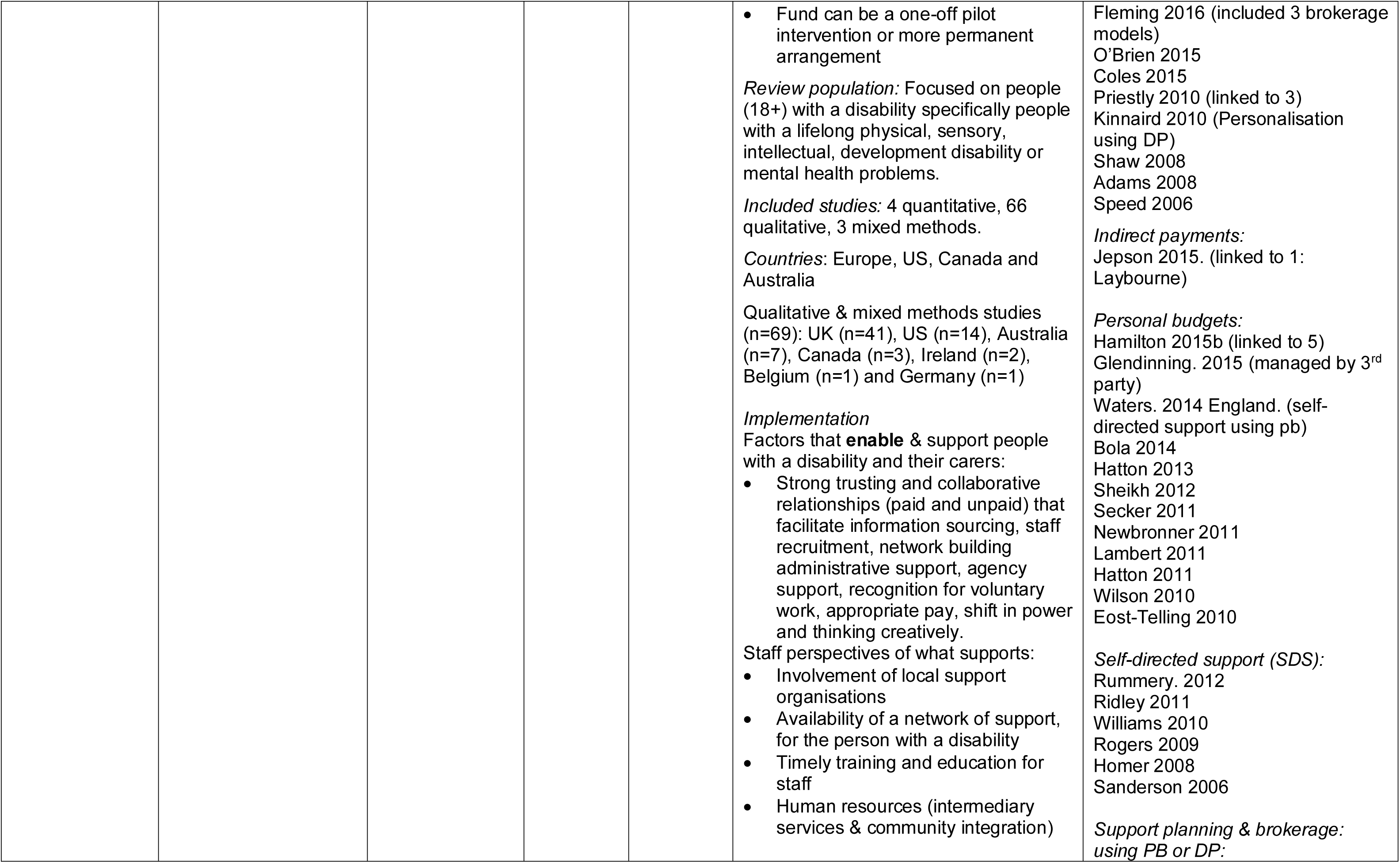
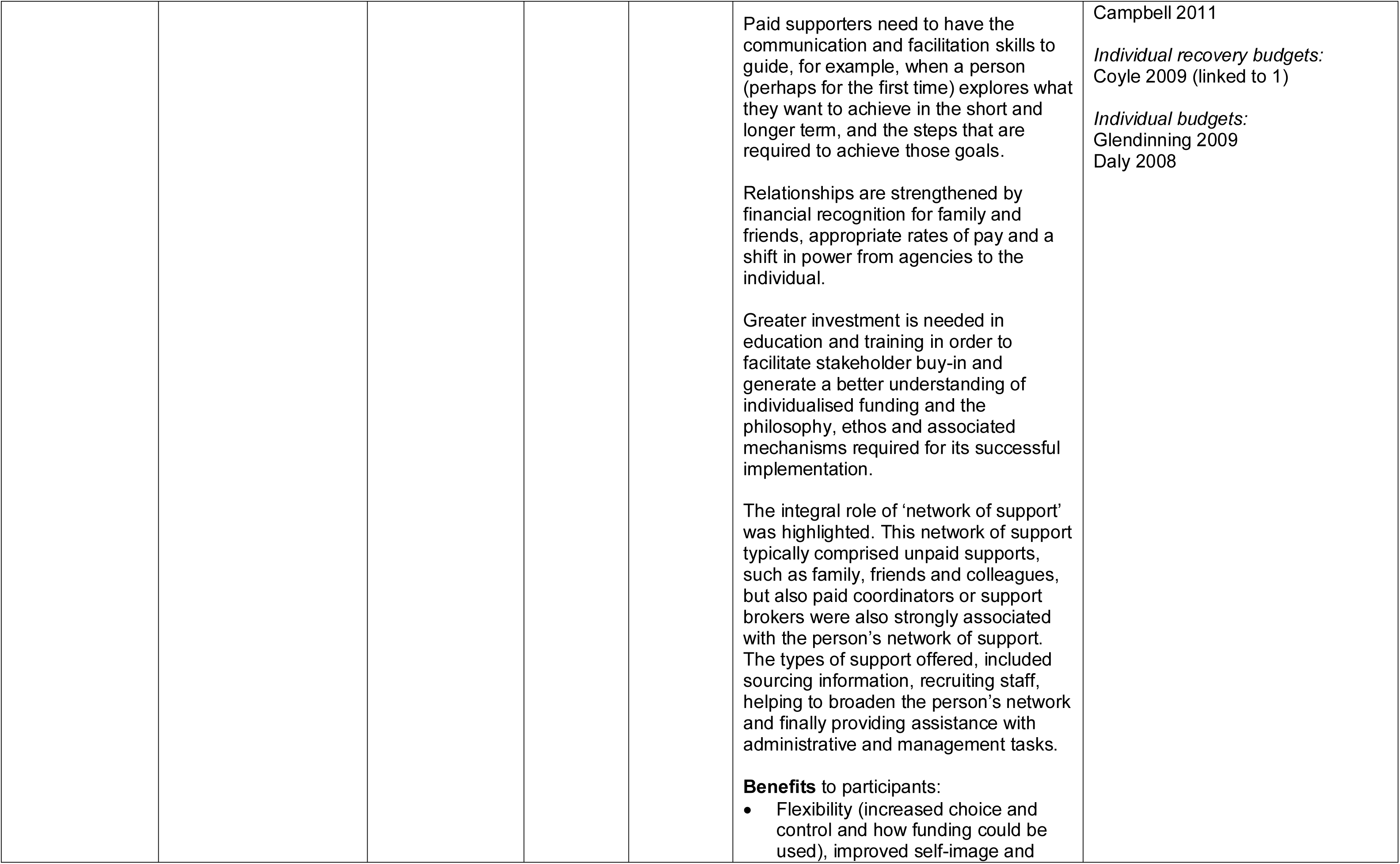
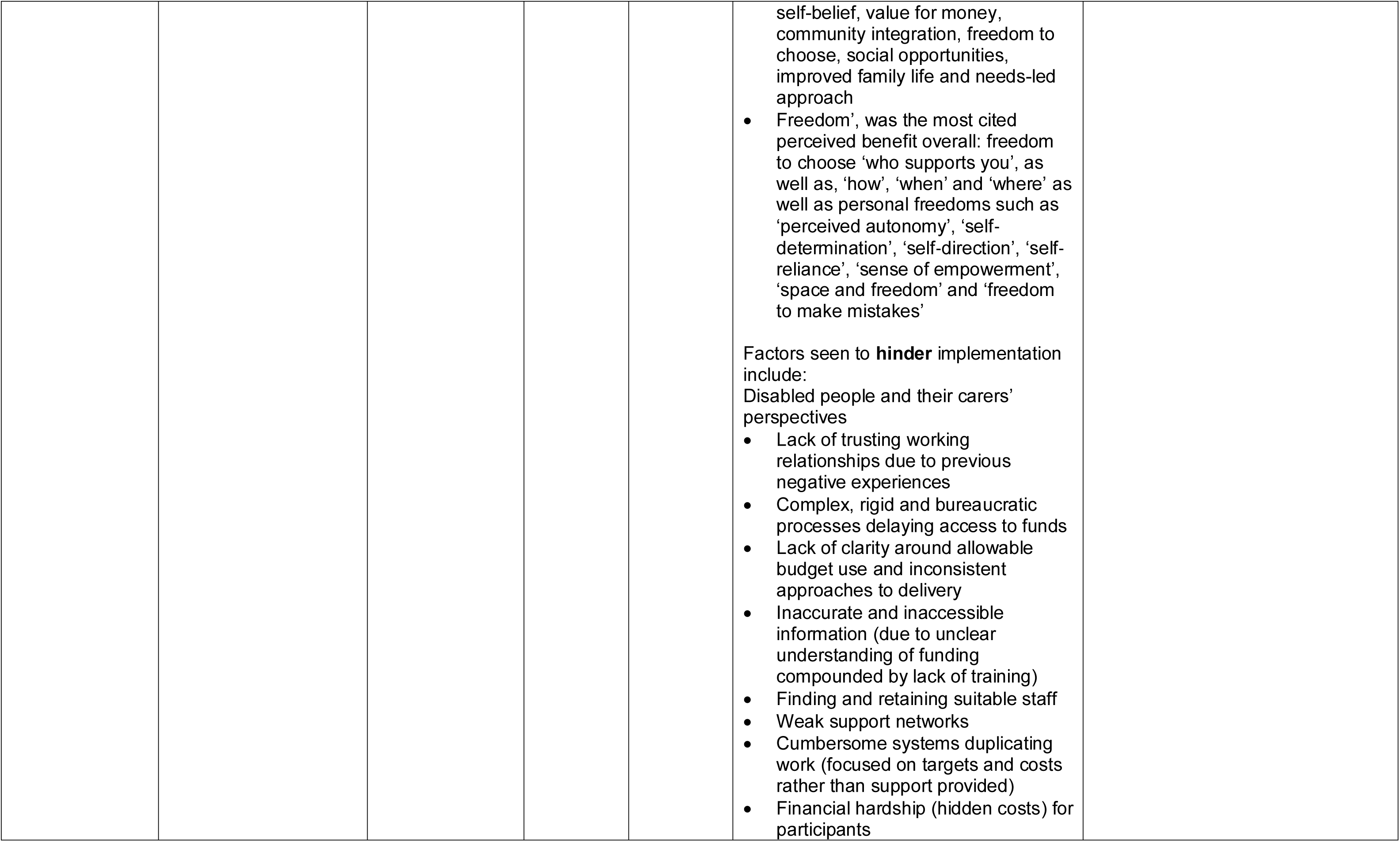
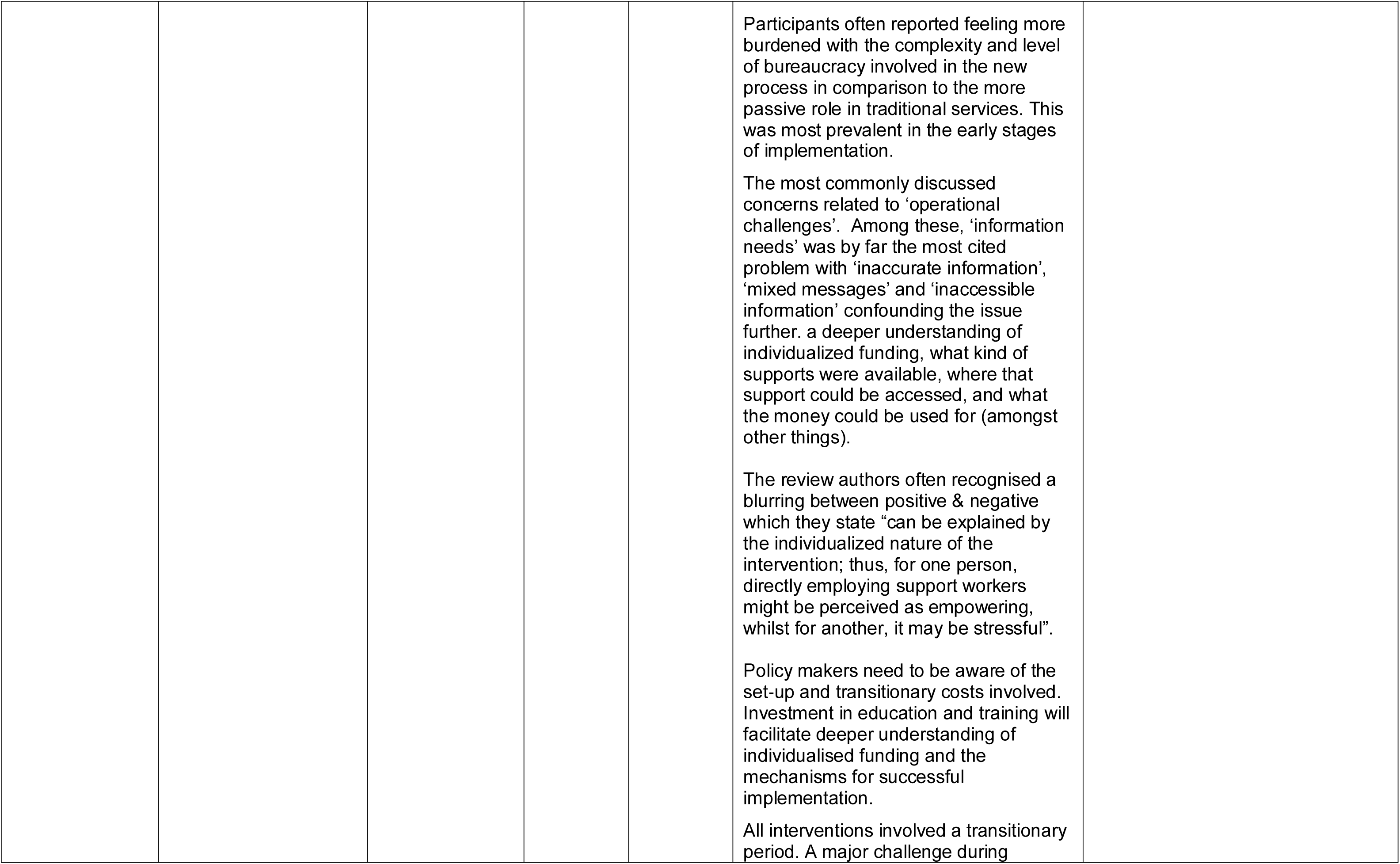
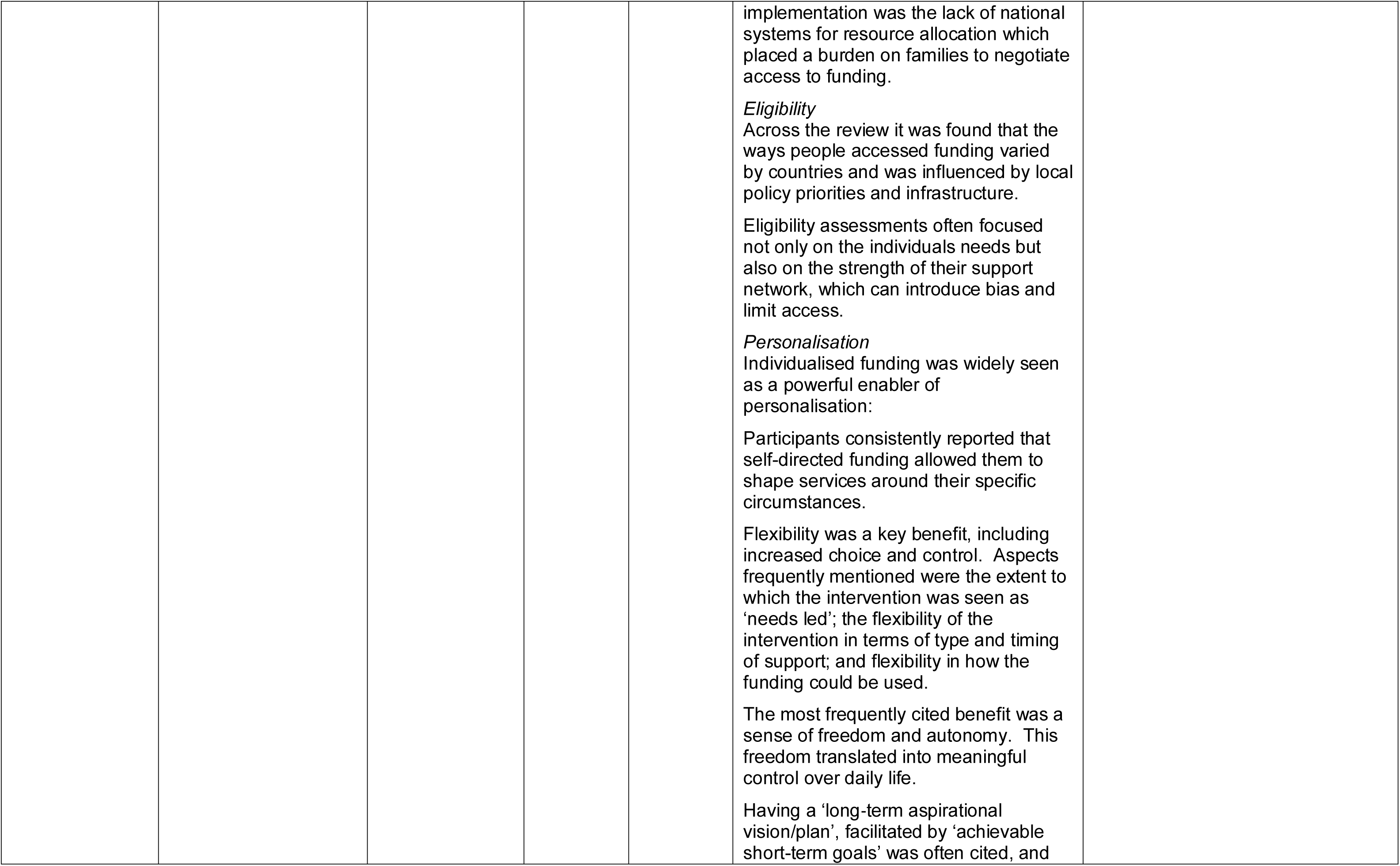
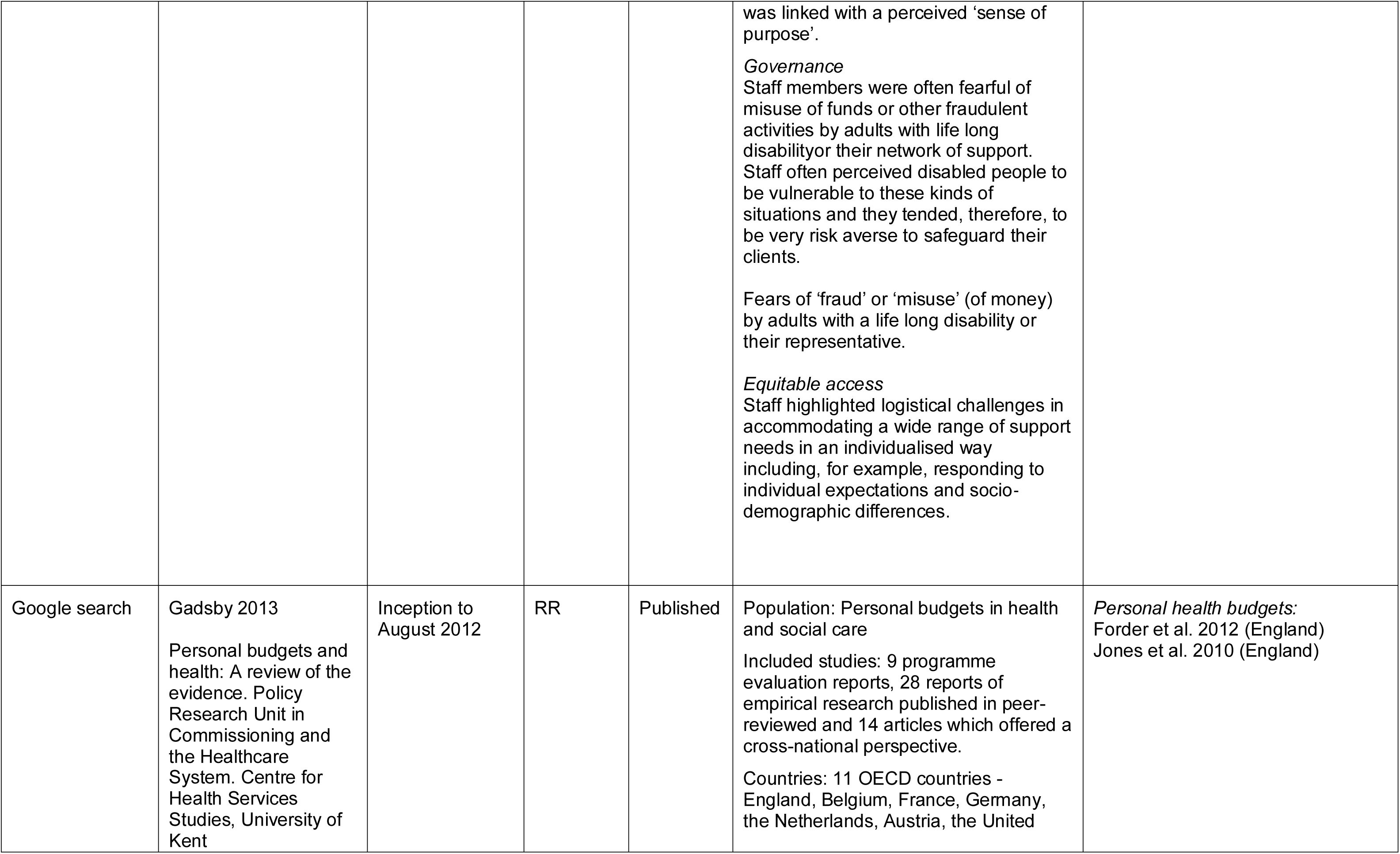
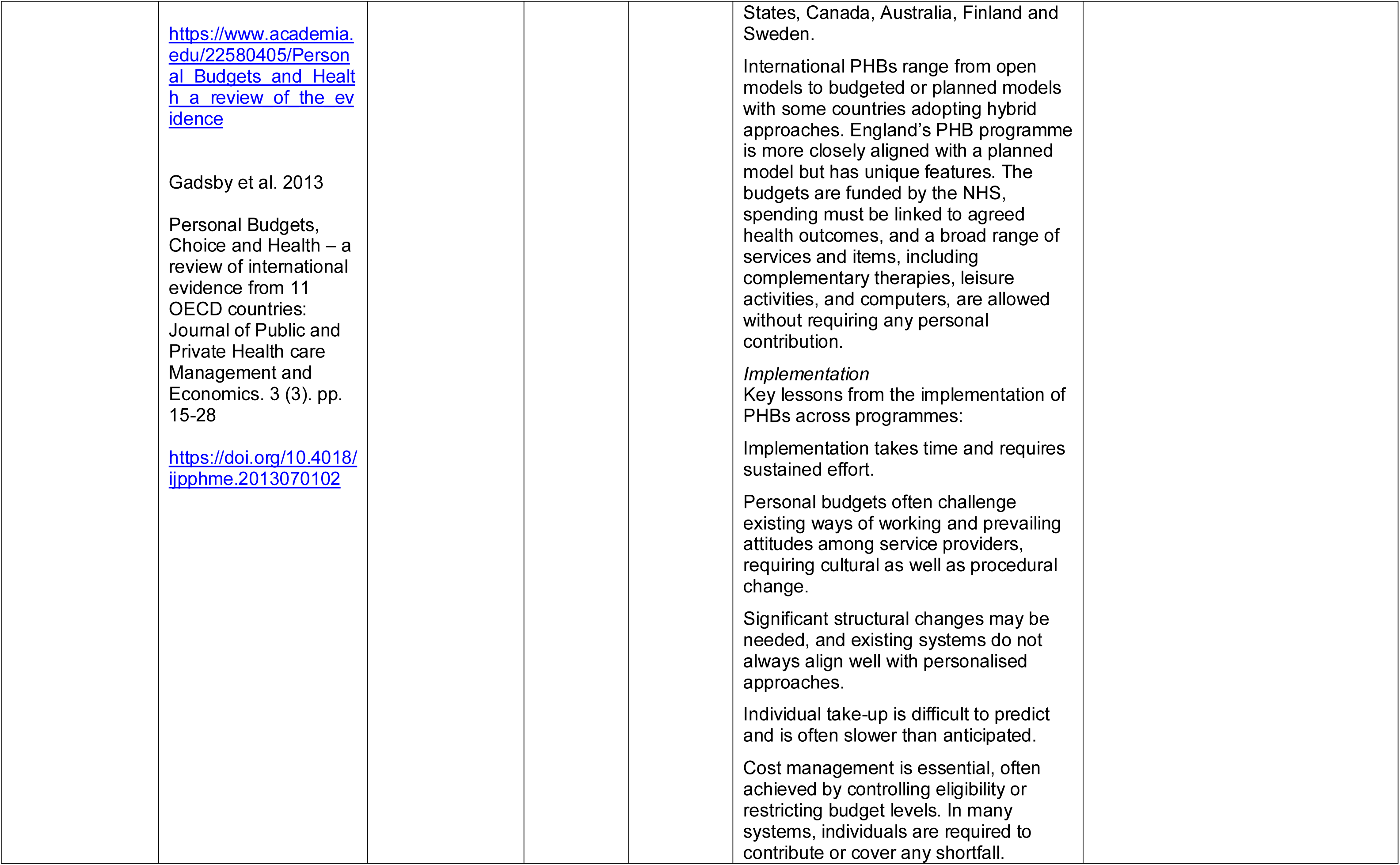
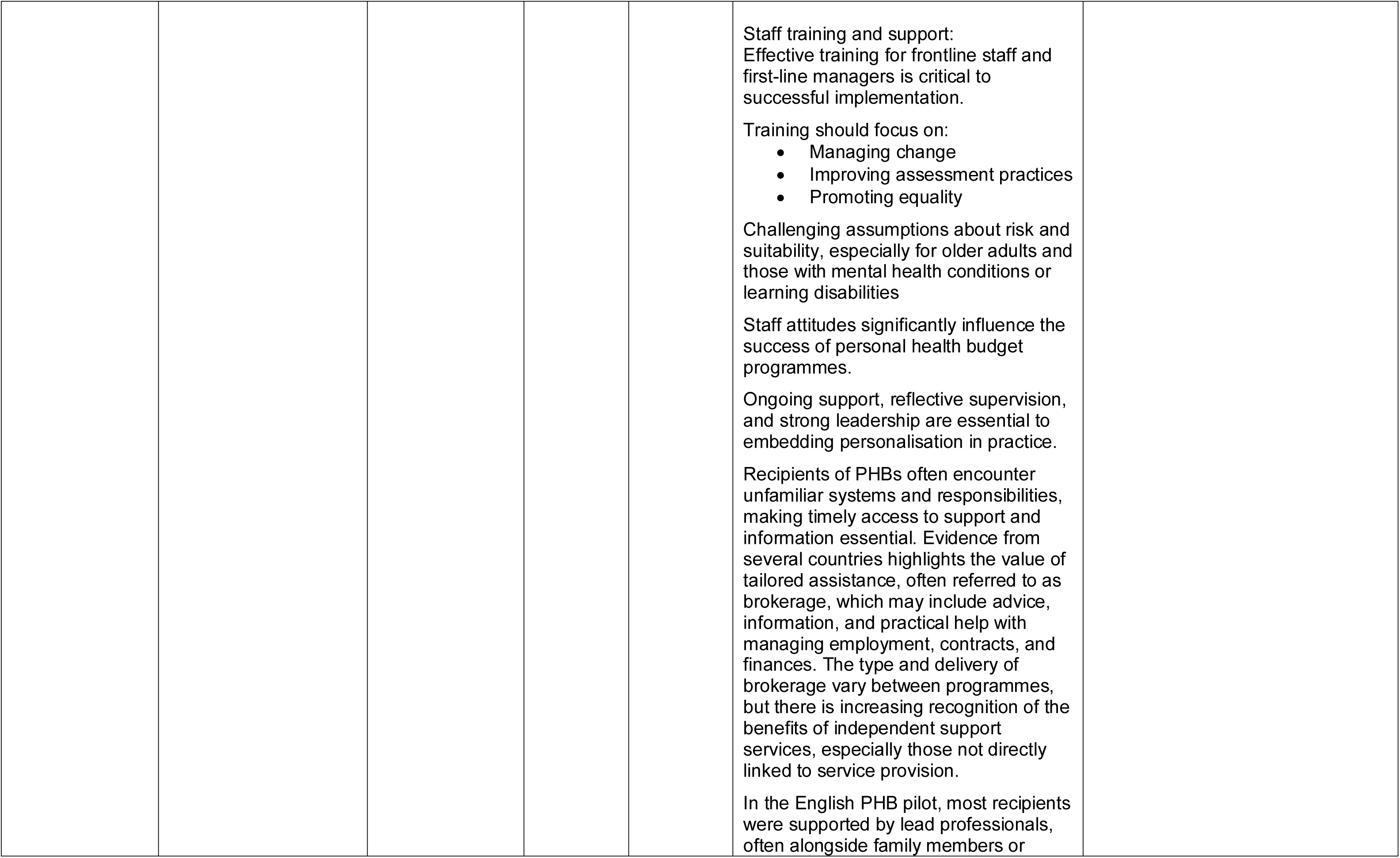
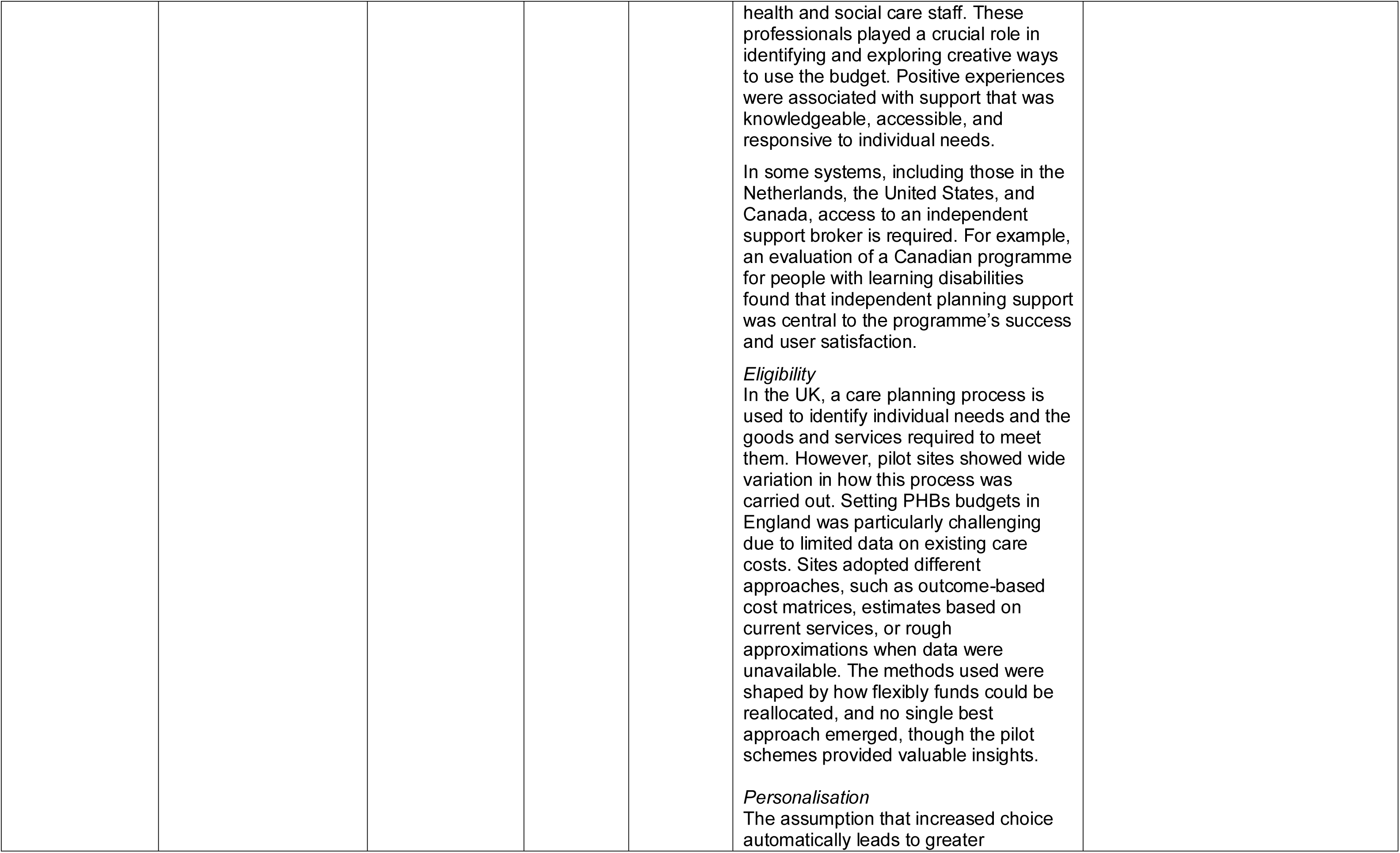
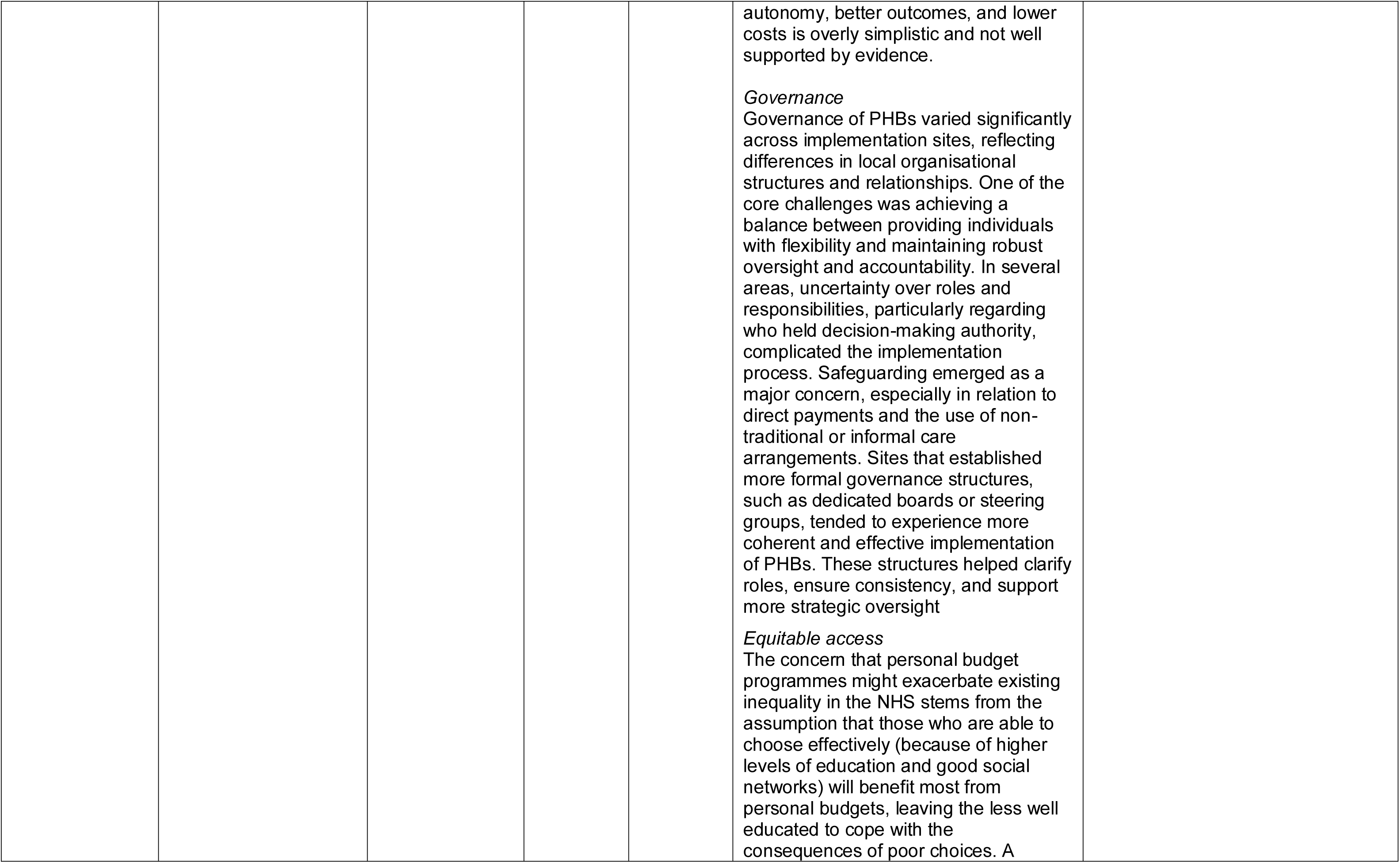
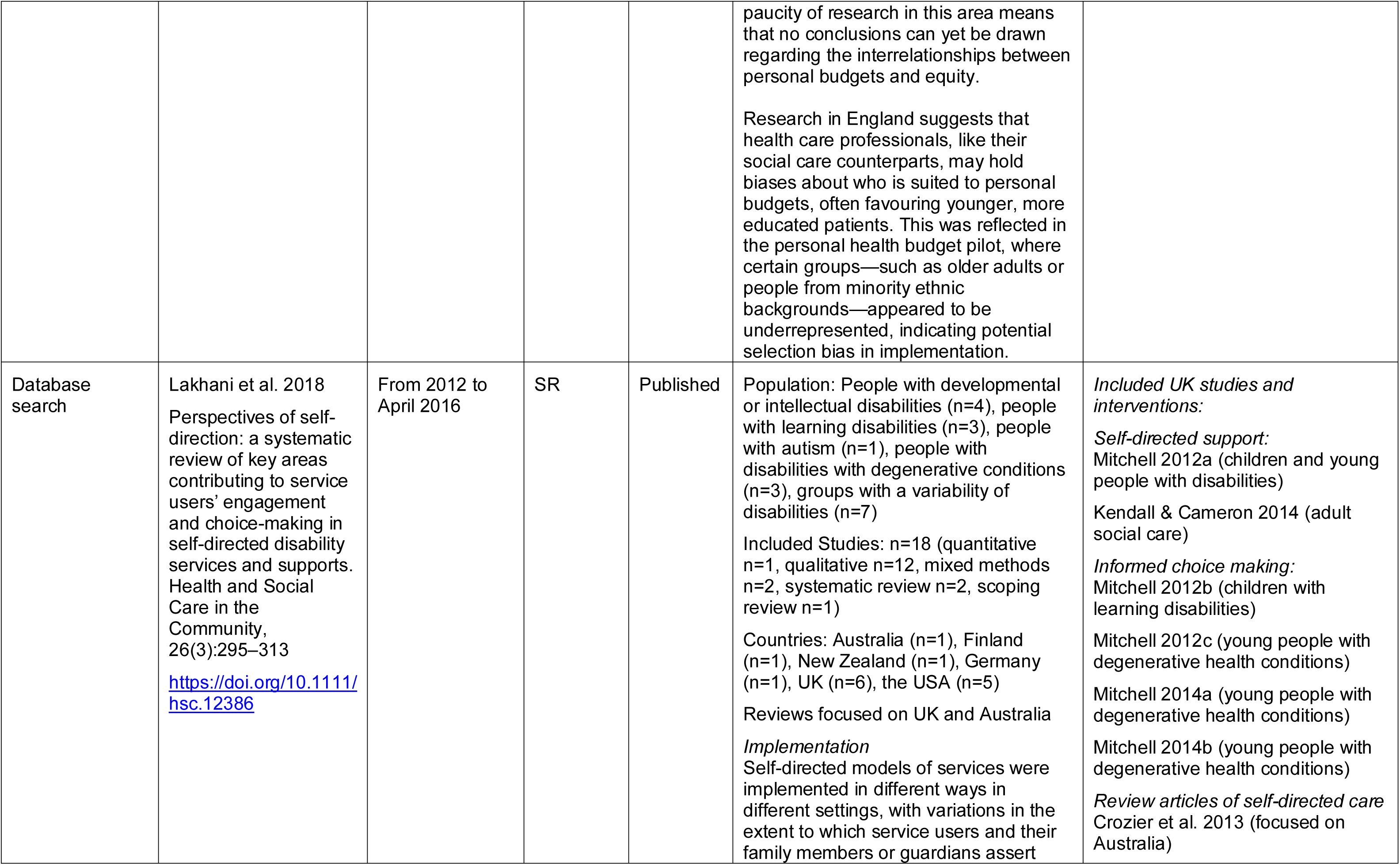
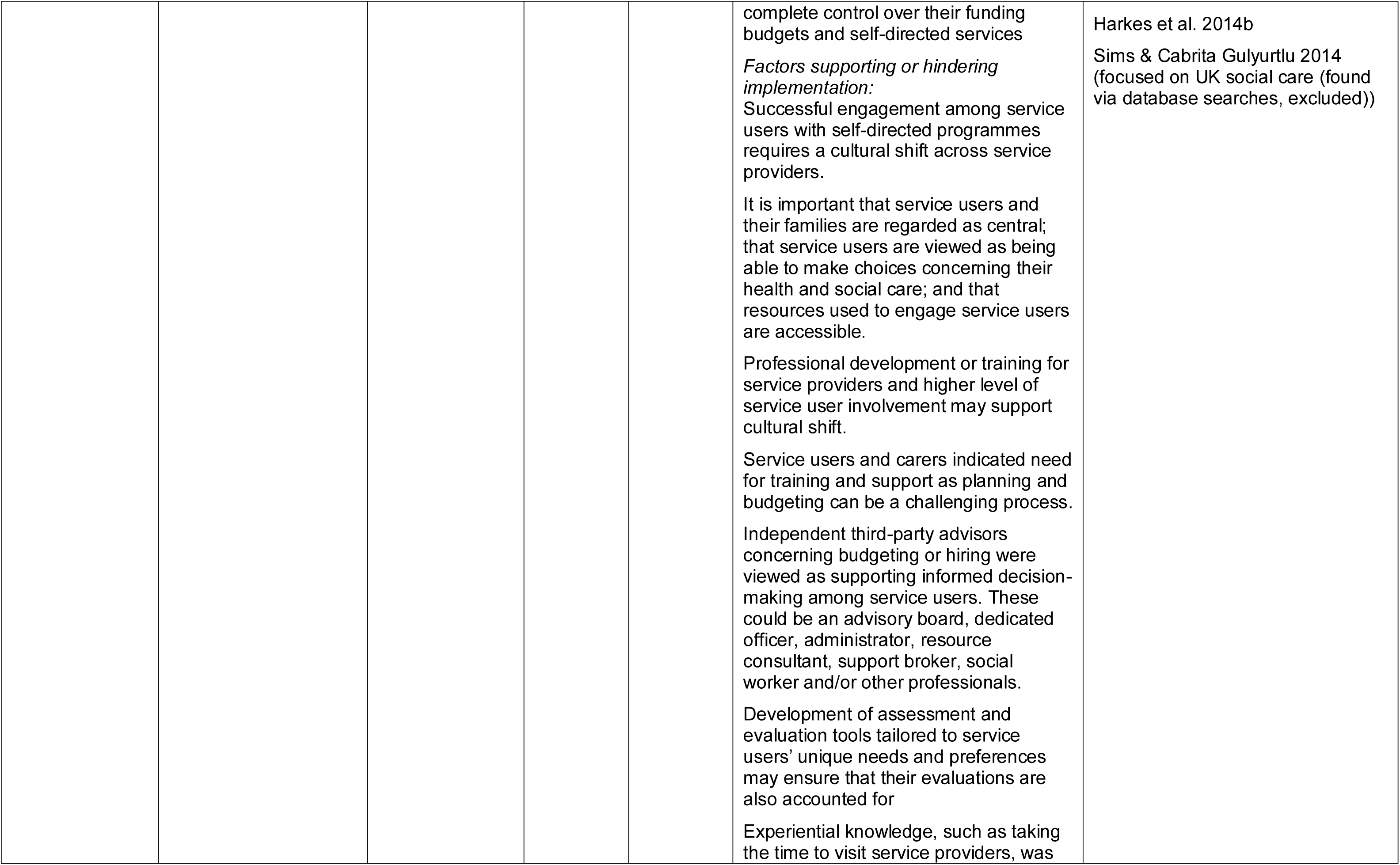
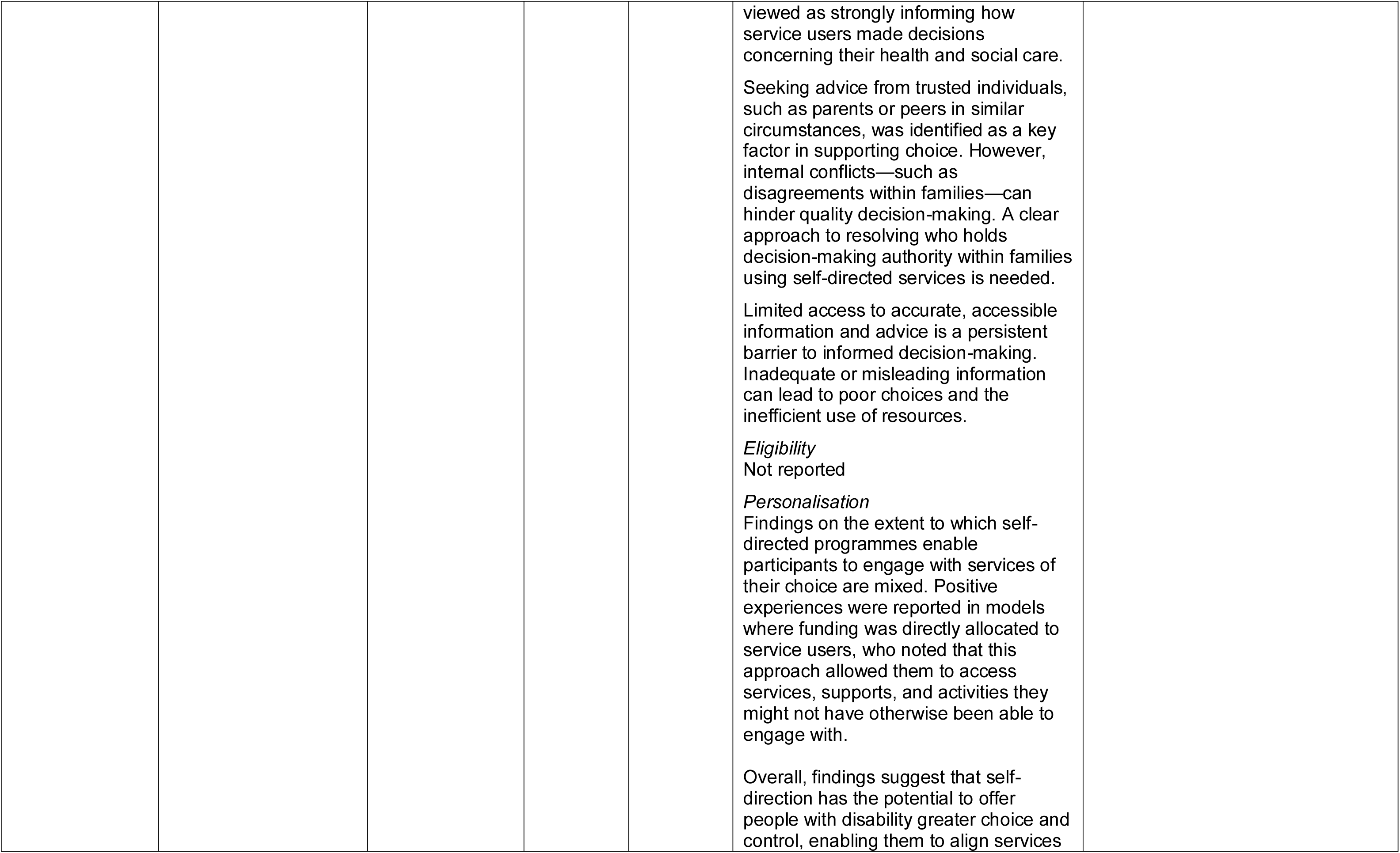
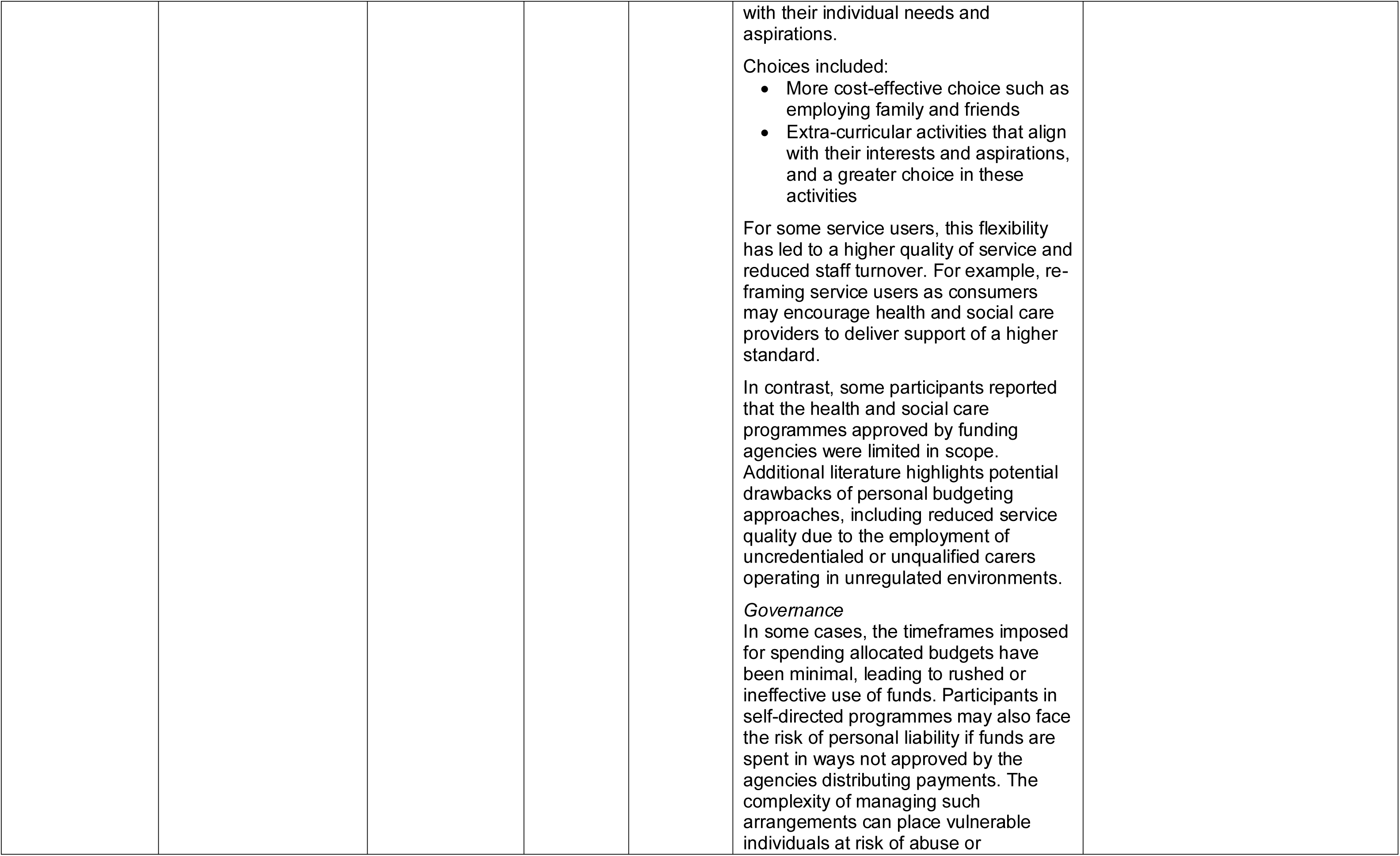
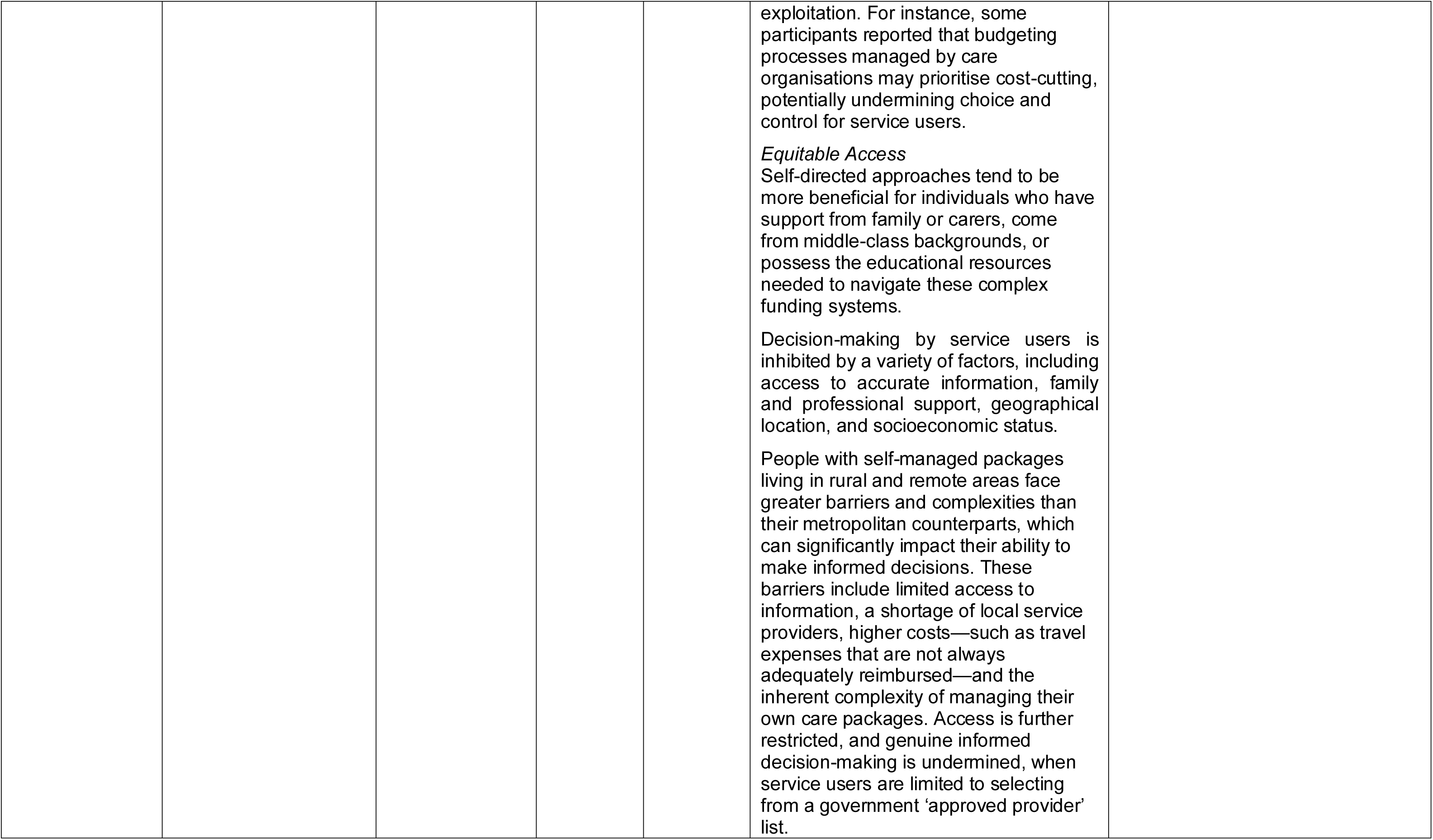
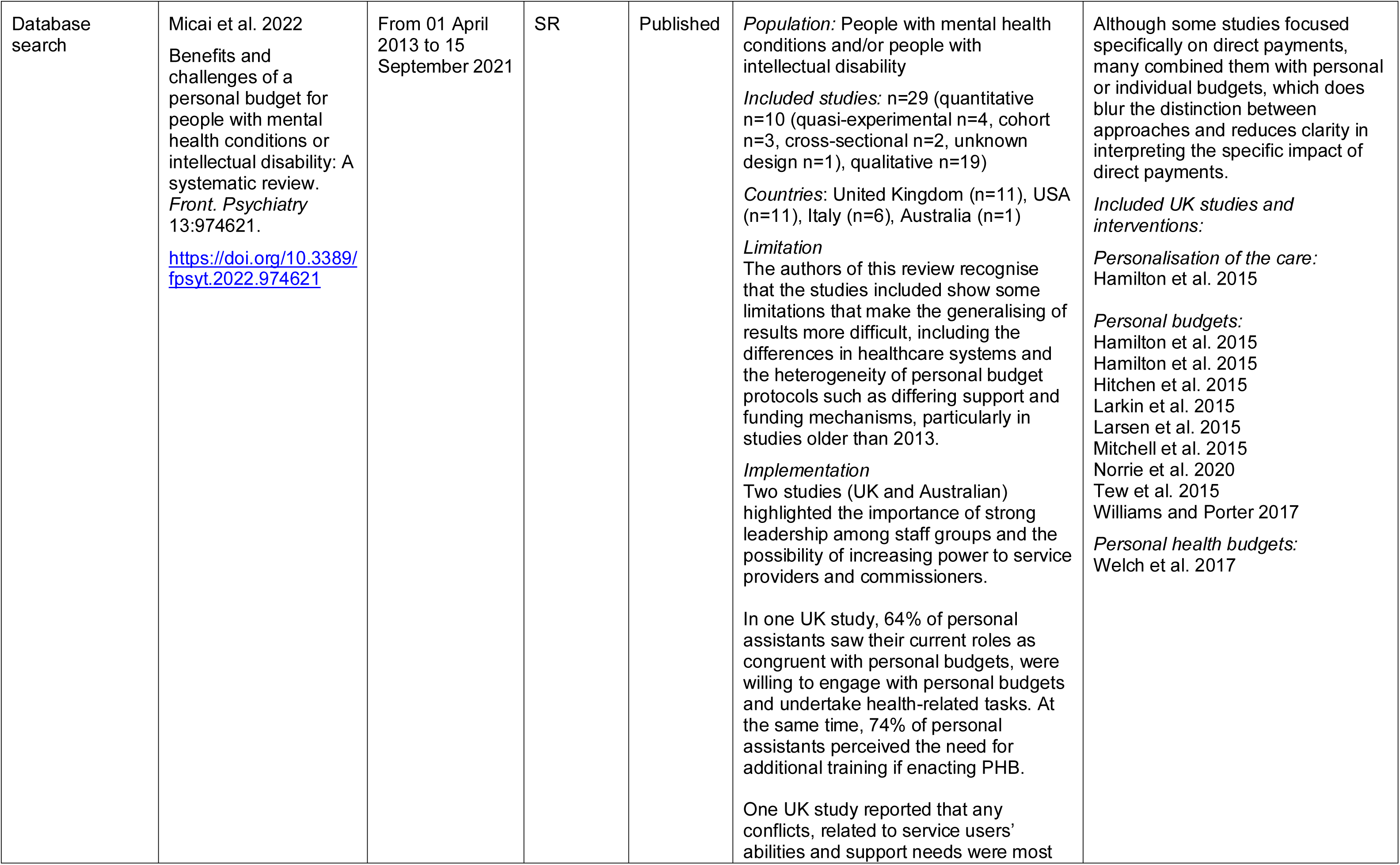
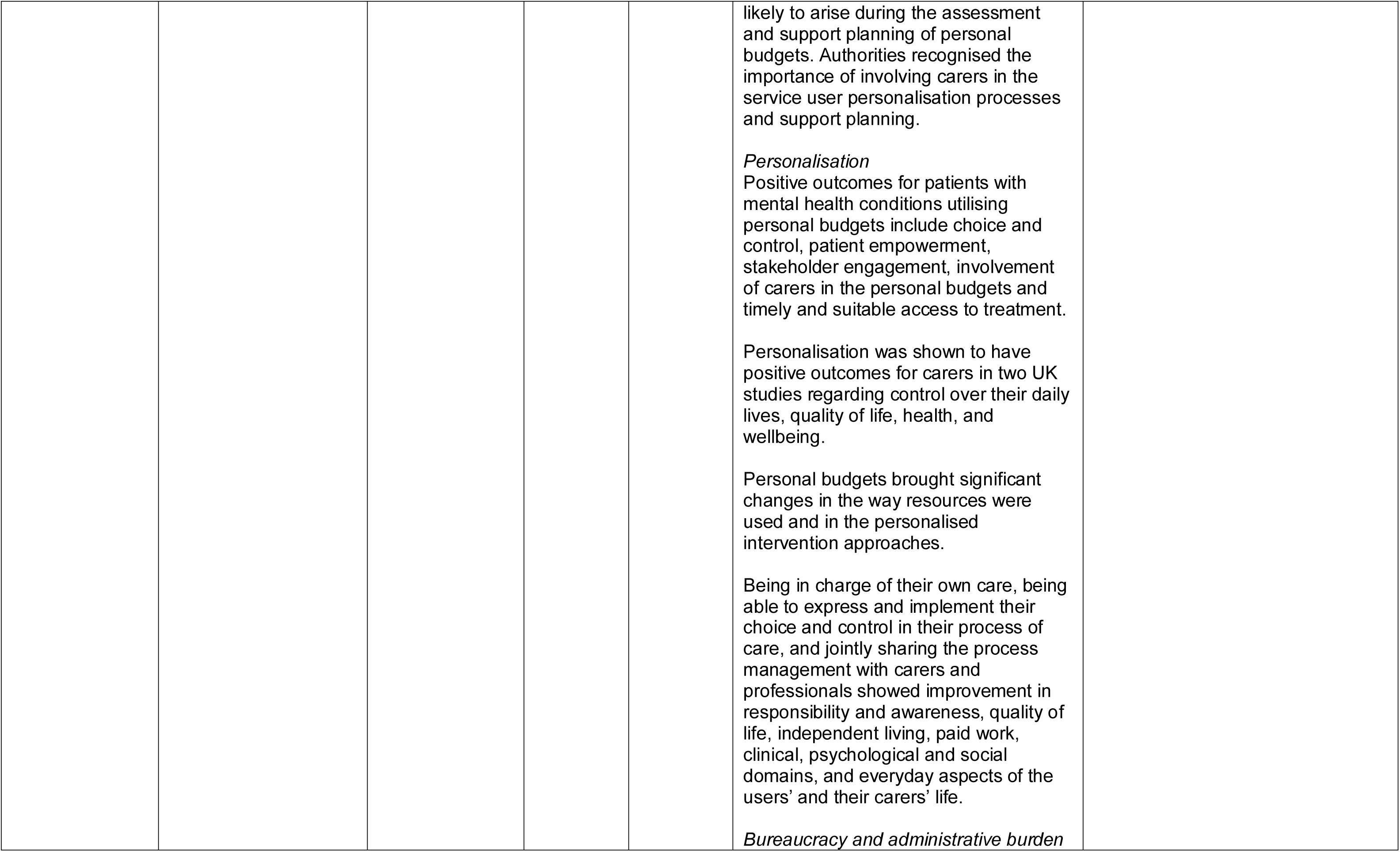
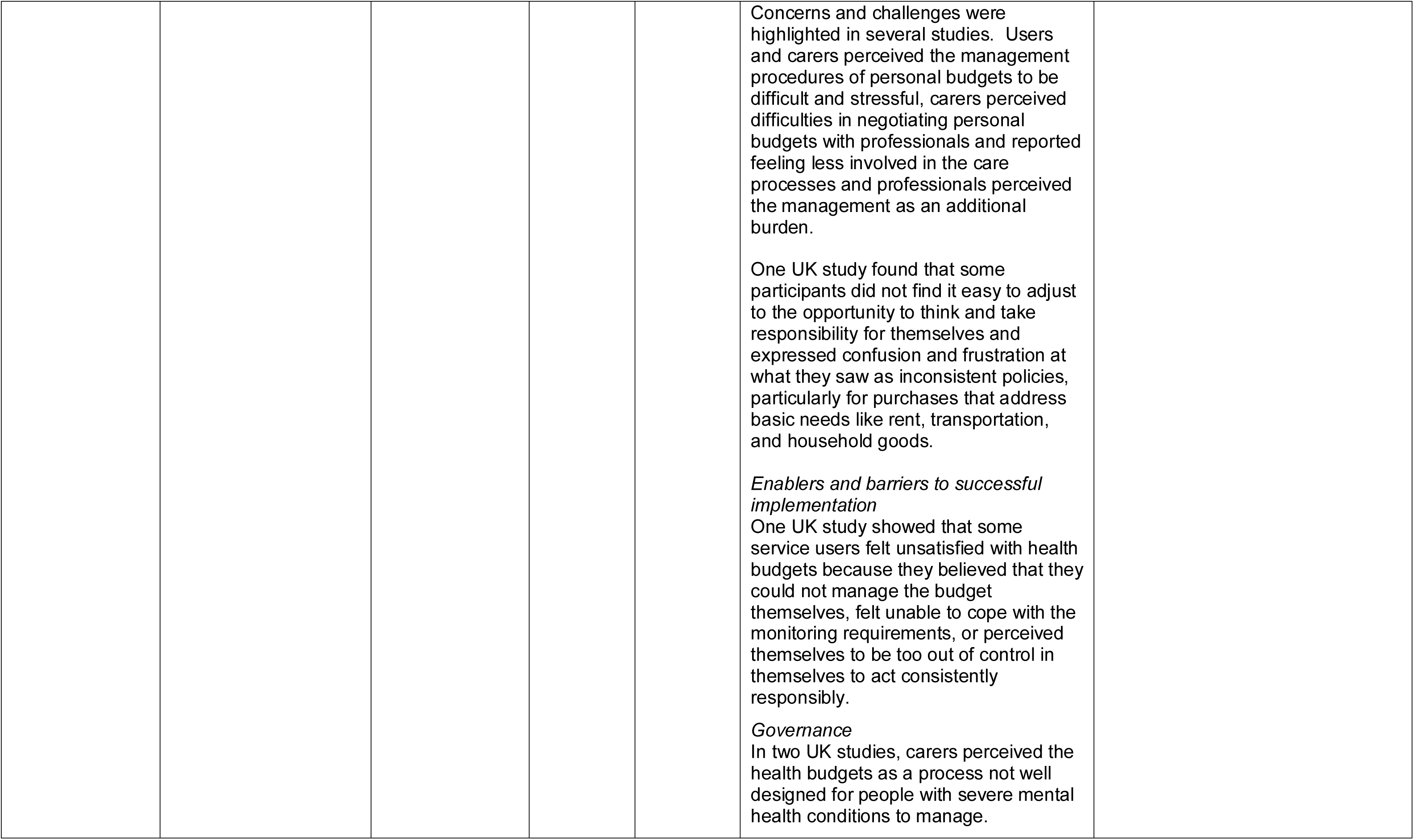
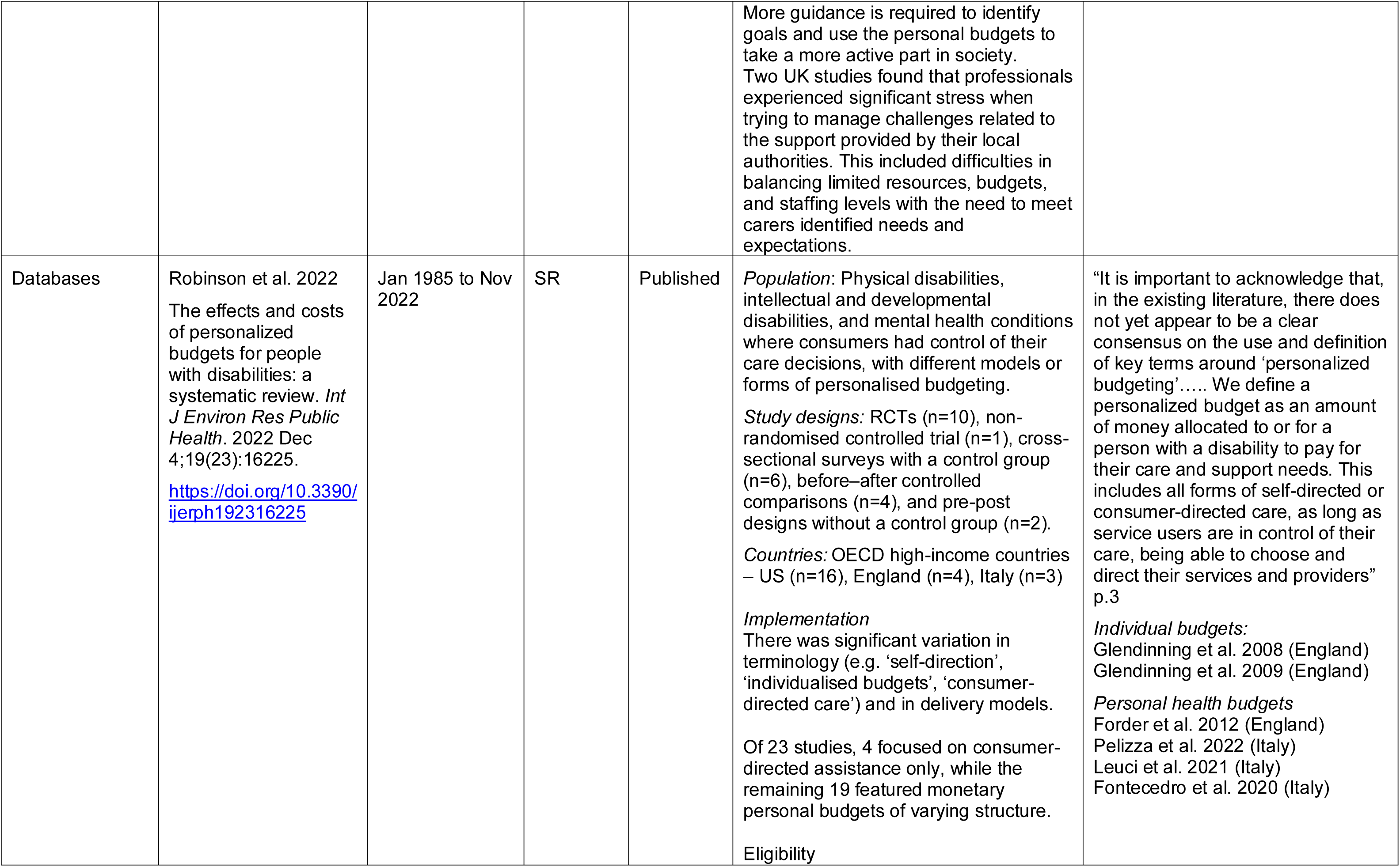
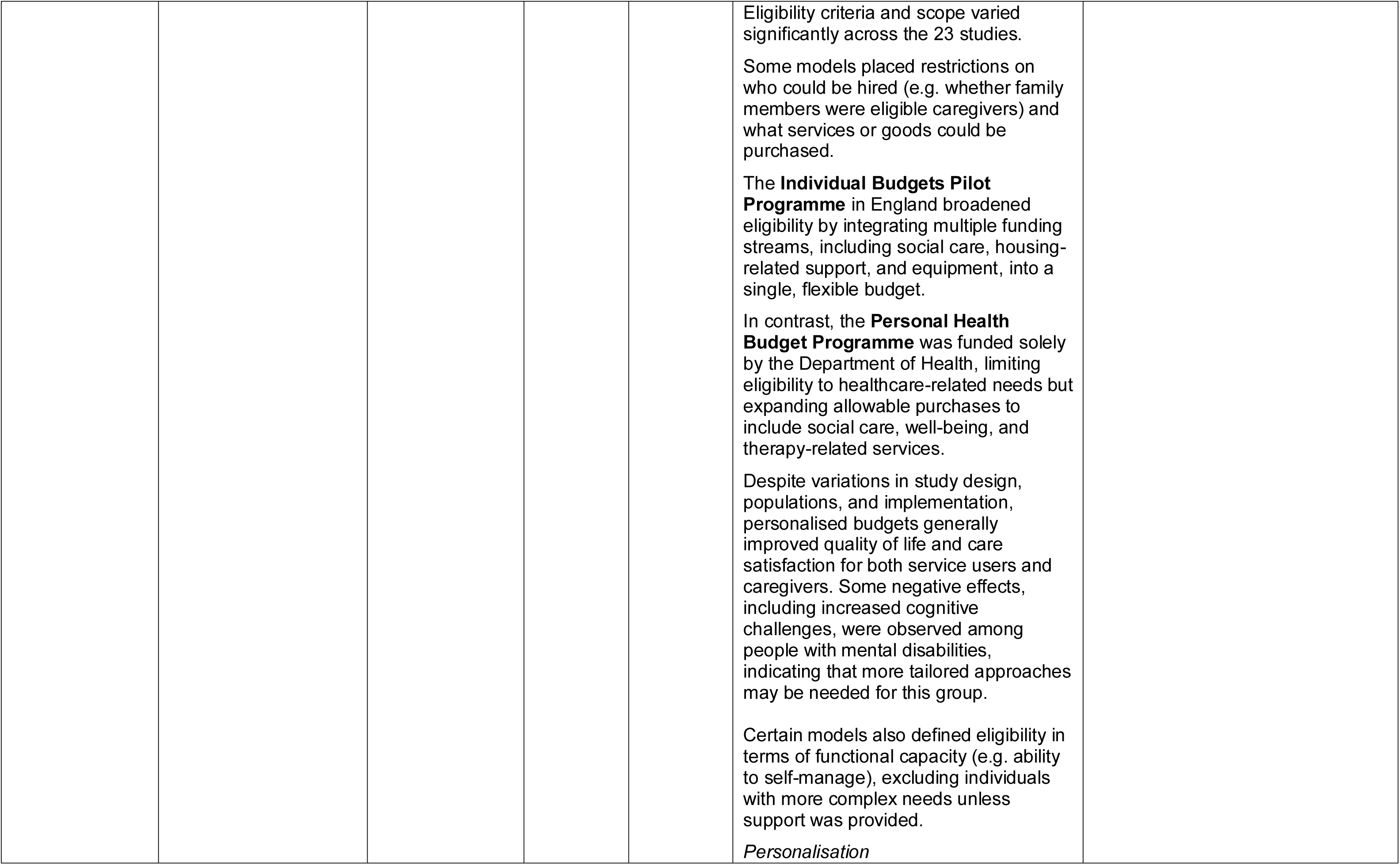
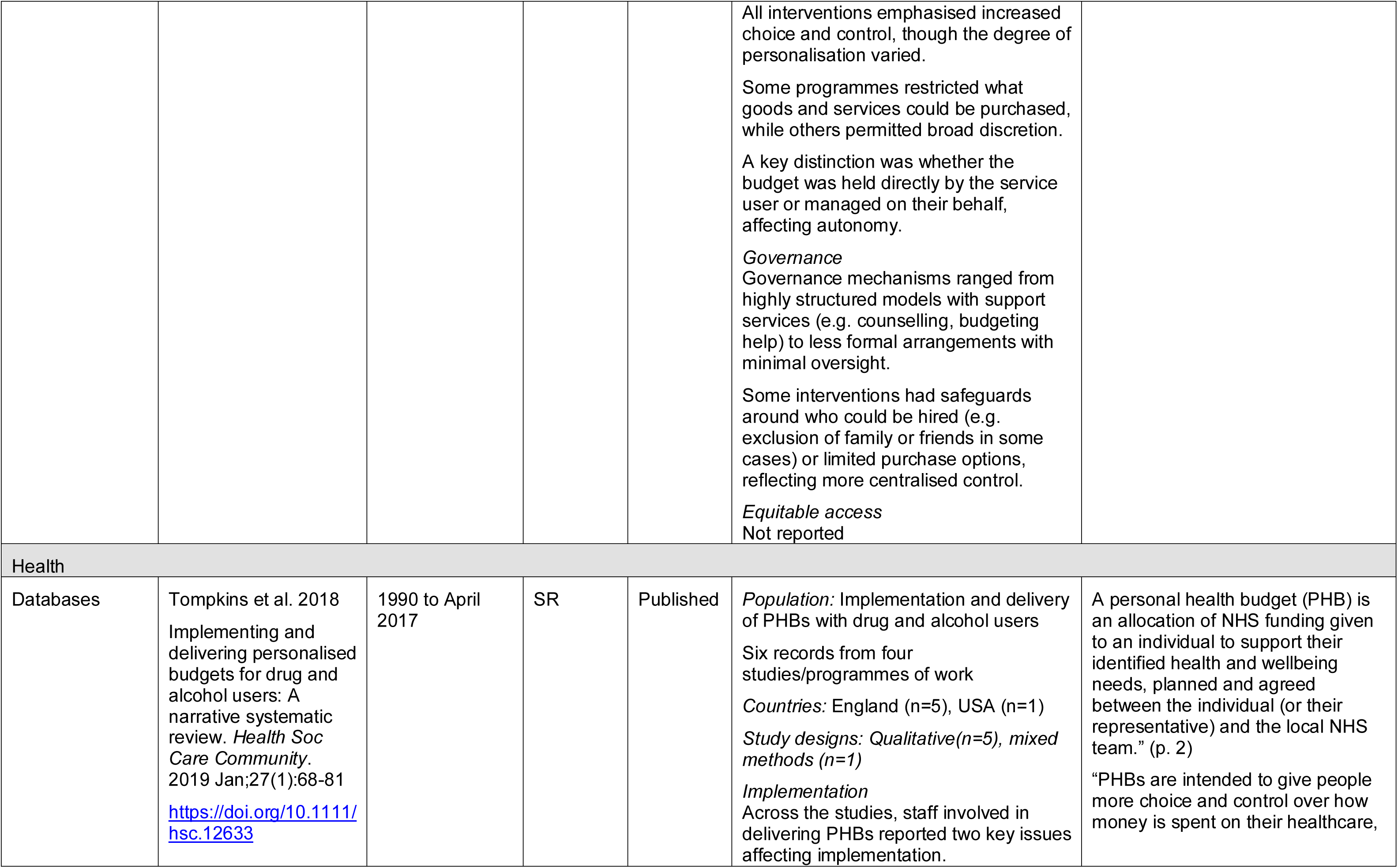
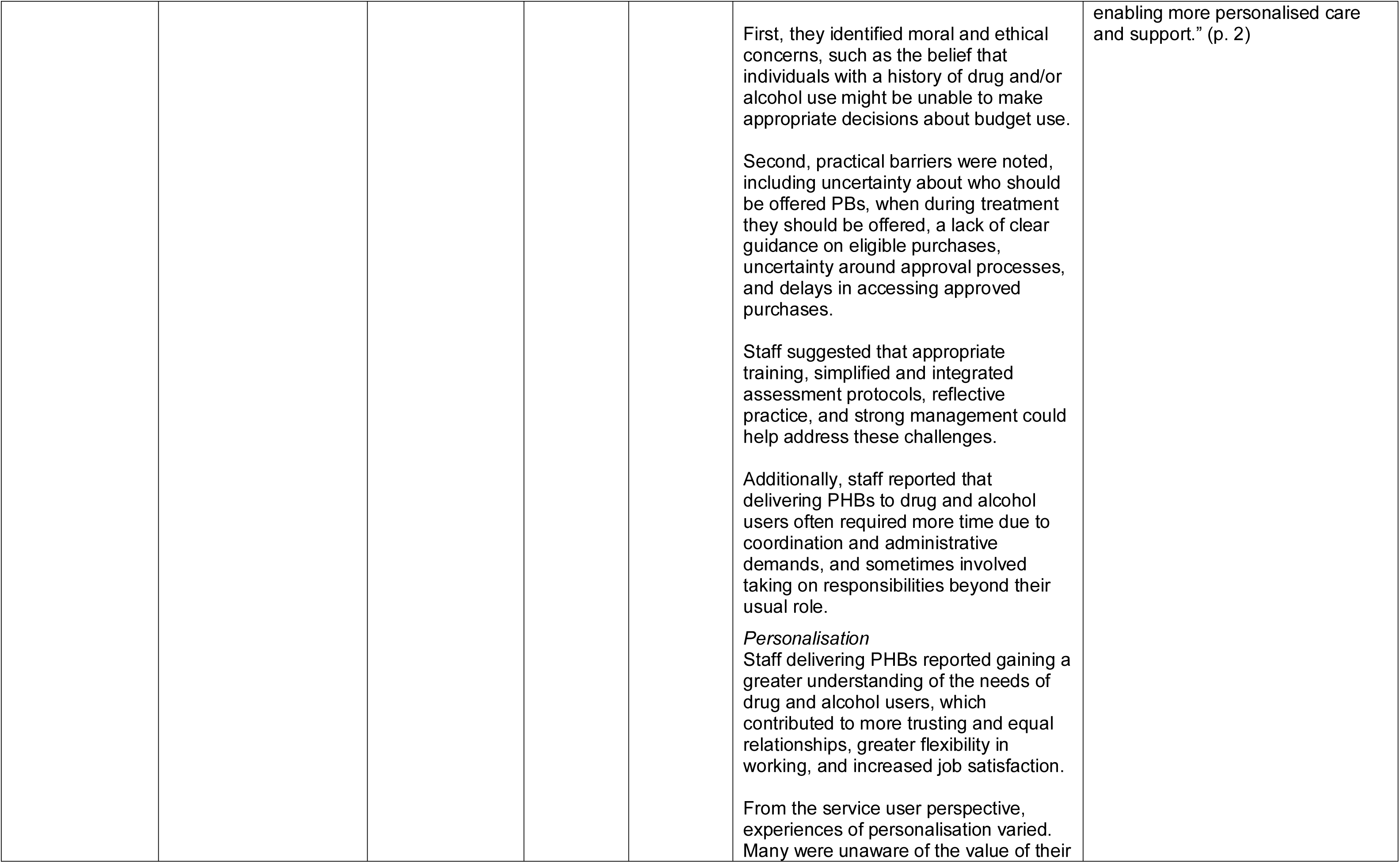
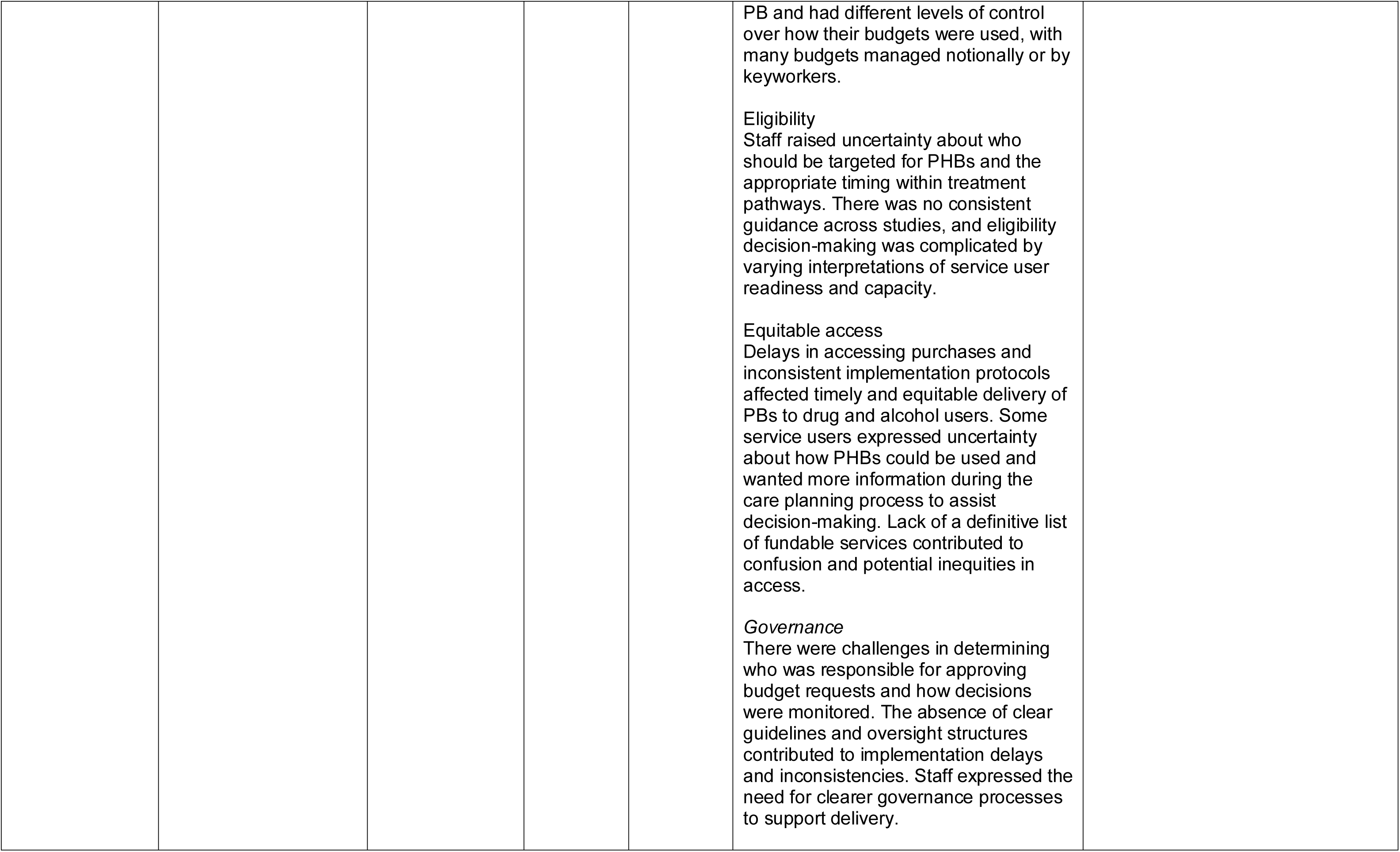
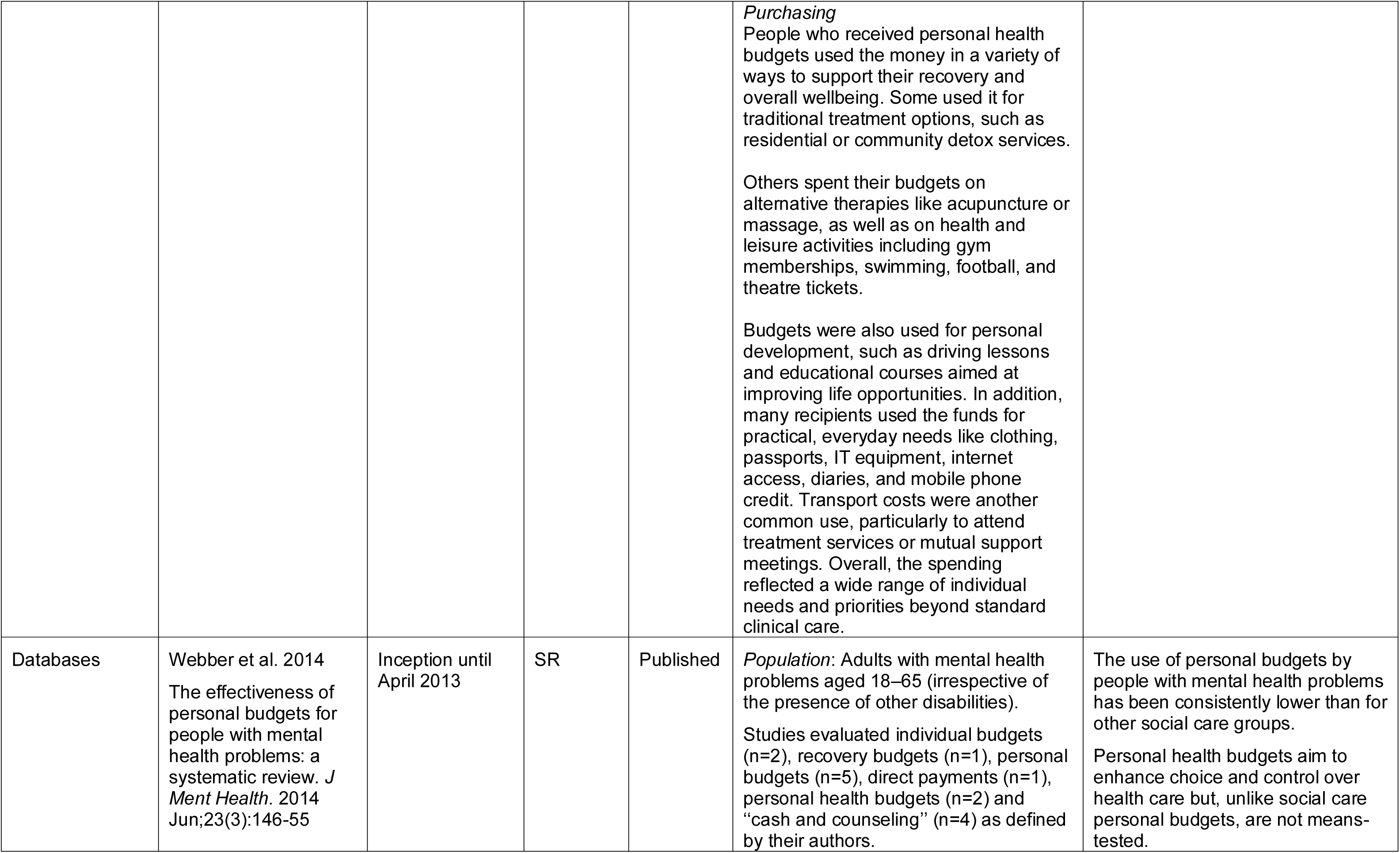
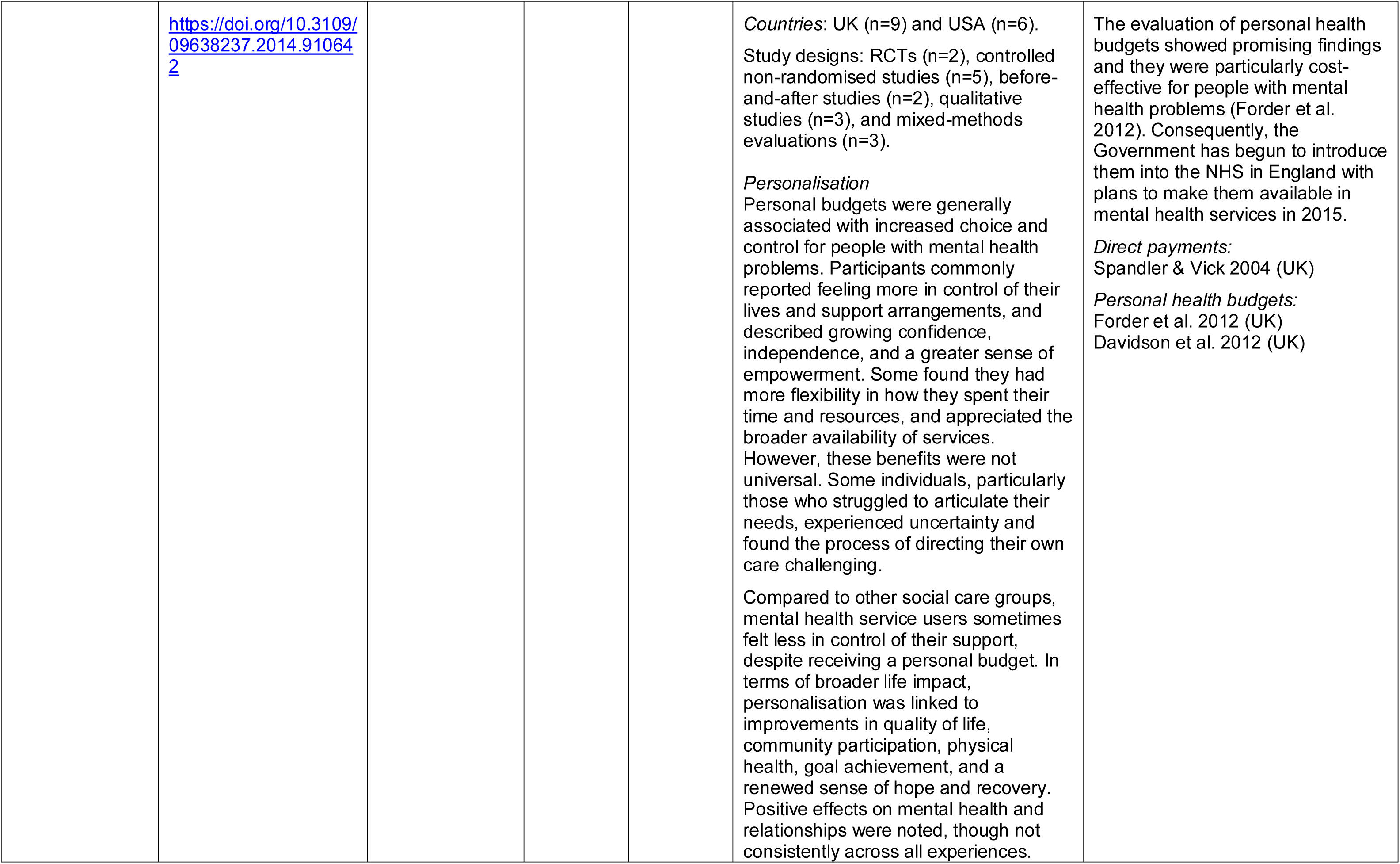
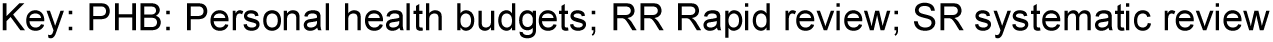
Summary of included review evidence.

**Table 3:** Summary of personal health budgets evaluation (pilot)

| **Citation** | **Content Summary** | **Population** | **Direct Payment Findings** |
| --- | --- | --- | --- |
| Jones et al. 2010a<br>Evaluation of the personal health budgets pilots. Outline of a research project funded by the Department of Health. Personal Social Services Research Unit, University of Kent.<br>https://www.pssru.ac.uk/publications/pub-1820/ | Project outline<br>Sets out the design and methodology for the national evaluation across 70 pilot sites with 20 selected for in-depth evaluation | Adults with CHC, long-term conditions, mental health needs, stroke survivors<br>Children with complex needs, represented by their parents or carers | Does not report findings as this is an outline and one of the aims was to explore the different mechanisms for delivering PHBs, including direct payments, notional budgets, and third-party arrangements. |
| Jones et al. 2010b<br>Early experiences of implementing personal health budgets. PSSRU Discussion Paper 2726/2. Personal Social Services Research Unit, University of Kent.<br>https://www.phbe.org.uk/documents/interim_report_july_2010.pdf | First interim report<br>Outlines the implementation progress and setup | Adults with CHC, long-term conditions, mental health needs, stroke survivors<br>Children with complex needs, represented by their parents or carers | The report confirms that direct payments were an intended delivery mechanism for PHBs, particularly for non-CHC groups.<br>Notes legal limitations on direct payments for CHC, which had not yet been resolved at this stage.<br>Mentions that some pilot sites were already using direct payments under social care legislation |
| Jones et al. 2010c.<br>Experiences of implementing personal health budgets: 2<sup>nd</sup> interim report. PSSRU Discussion Paper 2747/2. Personal Social Services Research Unit, University of Kent. | Second interim report<br>Presents early qualitative findings from the second wave of interviews, conducted between September and October 2010, across the 20 in-depth pilot sites. It explores how personal health budgets were | Interviews with commissioners, managers, and frontline staff.<br>Compared with the first two reports, it includes views from a wider range of staff, not just project leads, and offers a deeper exploration | *Factors that enable or hinder successful implementation*<br>Staff felt underprepared to support direct payments due to a lack of training and clear guidance.<br>*Personalisation*<br>While direct payments could offer more respite and flexibility, some carers were worried about increased responsibility and how it might affect their own lives and benefits. Direct payments were widely seen as the most empowering option, with many frontline staff and some |
| https://www.gov.uk/government/publications/experiences-of-implementing-personal-health-budgets-2nd-interim-report | being introduced, planned, and delivered | of the positive and negative impacts on carers | budget holders viewing them as the only true way to achieve full choice and control.<br>*Equitable access*<br>Direct payments were seen as having the potential to improve equity for Black and minority ethnic budget holders by enabling access to more culturally and linguistically appropriate support, including the option for family-led services.<br>*Governance*<br>Staff expressed anxieties about risk, especially when direct payments were used to employ family members or unqualified carers. There were concerns about fraud or misuse, such as spending the entire budget early or on non-health-related items. There was confusion about appeals processes and who is accountable if something goes wrong with a direct payment. |
| Jones et al. 2011.<br>The cost of implementing personal health budgets. Personal Social Services Research Unit, University of Kent.<br>https://www.gov.uk/government/publications/the-cost-of-implementing-personal-health-budgets | Third interim report<br>Examines the financial costs associated with planning and delivering PHBs through different models across 20 of the pilot sites | Adults with CHC, long-term conditions, mental health needs, stroke survivors<br>Children with complex needs, represented by their parents or carers | Four pilot sites reported spending on setting up direct payment services. |
| Irvine et al. 2011<br>Personal health budgets: early experiences of budget holders, Fourth Interim Report. DH 2478. Personal Social Services Research Unit, University of Kent | Fourth interim report<br>Presents findings from in-depth interviews with 58 personal health budget holders (or their representatives) across 17 pilot sites.. | Budget holders, i.e., individuals who had already been allocated a PHB<br>People with long-term physical health conditions<br>Individuals with mental health needs<br>Parents of children with complex needs | Direct payments are mentioned throughout as one of the key mechanisms by which individuals accessed their PHB.<br>*Factors that enable or hinder successful implementation*<br>Motivations for choosing direct payments included because they suited their lifestyle, offered greater choice and flexibility, or avoided third-party fees. Others had prior experience or enjoyed the admin.<br>The stress of managing care was already significant for some, leading a few to use payroll services to reduce the administrative burden. Details of the direct payment option were not always clearly explained, such as whether the full annual budget would be paid upfront. Clear |
| https://www.gov.uk/government/publications/personal-health-budgets-fourth-interim-report |  |  | information about the direct payment option is needed both during and after the care planning process, as some questions may only arise once the personal health budget is in use. |
| Davidson et al. 2012<br>Personal health budgets: experiences and outcomes for budget holders at nine months, Fifth interim report. DH 2523. Personal Social Services Research Unit, University of Kent.<br>https://www.gov.uk/government/publications/personal-health-budget-pilots-fifth-interim-evaluation-report | Firth interim report<br>Explores what budget holders actually did with their PHBs, once implemented<br>Insights into purchases made<br>How budgets were managed (e.g. direct payments, third-party)<br>Day-to-day outcomes and challenges | Carers of adults receiving PHBs across various pilot sites and health conditions | *Factors that enable or hinder successful implementation*<br>Nearly half of those using direct payments reported initial issues, such as setting up bank accounts, managing payments, and delays in funds being deposited or notified. Some struggled with drafting contracts and used online templates.<br>Most people using direct payments felt it was the right option.<br>About half of budget holders employing staff managed employment tasks like tax and National Insurance themselves, while the other half used an agency. Those managing it alone (employment tasks) often felt daunted and relied on support from professionals, friends, or family. Some did not need recruitment support, having retained staff from previous care packages or using agency carers. Support from PHB lead officers or support workers was highly valued, including help with advertising, shortlisting, and conducting interviews.<br>Many employed their own carer or personal assistant, often friends, relatives, or previous care staff. Holding interviews in professional settings rather than at home was described as a significant relief. - Some faced difficulties recruiting suitable staff locally. Challenges included the cost of advertising and finding applicants willing to work on payroll rather than cash in hand. A few experienced problems when existing care arrangements broke down. Some budget holders did not feel CRB checks were necessary, particularly when employing relatives or staff who already had checks in place. Others found support with arranging CRB checks, such as through a support worker to be very helpful. A few faced difficulties with staff, and in one case, a support worker helped dismiss a carer, highlighting the value of back-up support in managing employment issues. |
| Forder et al. 2012.<br>Evaluation of the Personal Health Budget Pilot Programme. PSSRU Discussion | The final report of the PHB evaluation, using a three-year mixed-methods approach to assess outcomes, service use, and cost-effectiveness across different health groups | Adults with CHC, long-term conditions, mental health needs, stroke survivors | Just under 45% of personal health budgets in the study were deployed as direct payments (as opposed to notional budgets or using third parties).<br>Personalisation |
| Paper 2840_2. Personal Social Services Research Unit, University of Kent.<br>https://www.york.ac.uk/inst/spru/research/pdf/phbe.pdf | It involved 2,700 participants, with 2,235 in the active sample<br>The study used a controlled trial design with both quantitative and qualitative data over 12 months<br>Qualitative interviews with PHB holders, carers, and professionals (overlap with the findings of the fifth interim report from Davidson et al. 2012) | Children with complex needs, represented by their parents or carers | Over two-thirds of carers (68%) reported a positive impact on their well-being when the PHB was delivered through direct payments.<br>The results suggest that the use of a PHB has a direct impact on quality of life via improved choice, control and tailoring of services to personal needs and circumstances.<br>*Factors that enable or hinder successful implementation*<br>Those with direct payments generally felt it was the right choice, despite common initial issues such as setting up bank accounts and delays in PCT payments.<br>Most participants using direct payments employed their own carers or personal assistants. Some required no recruitment help, having existing social care-funded carers or employing friends or family. Others, particularly first-time recruiters, valued support from PCT staff or support workers with tasks like advertising, interviewing, CRB checks, and contracts—relieving significant pressure.<br>However, challenges remained, including difficulty finding suitable carers willing to work on a payroll rather than a cash-in-hand basis. Ongoing support for managing employment arrangements was also seen as essential.<br>**Recommendation: Procurement and recruitment support for personal health budget holders (especially direct payment options) are valued by recipients.** |
| Jones et al. 2013<br>Personalisation in the health care system: do personal health budgets have an impact on outcomes and cost? Journal of Health Services Research & Policy, 18(2 Suppl), 59–67.<br>https://doi.org/10.1177/1355819613503152 | Summarises key evaluation results with some additional quantitative analysis and outcome data | Adults with CHC, long-term conditions, mental health needs, stroke survivors<br>Children with complex needs, represented by their parents or carers | Overlaps with Forder et al. 2012 and is reported within Fleming et al. 2019 |
| Welch et al. 2013<br>Implementing personal health budgets within substance misuse services. Final report. PSSRU Discussion Paper 2858. Personal Social Services Research Unit, University of Kent.<br>https://www.phbe.org.uk/documents/substance_misuse_final_report.pdf | Focused evaluation in two of the sites on people with substance misuse issues, reporting on impact, satisfaction, relapse, and implementation challenges in using PHBs | Adults with substance misuse issues<br>The majority of care/support plans were managed notionally. While one of the pilot sites did have approval to offer direct payments, we did not find evidence this deployment was offered during the pilot programme<br>The in-depth interviews with organisational representatives explored the various implementation issues and challenges around offering the direct payment deployment option to this client group | *Personalisation*<br>There was broad support for the potential of direct payments to increase flexibility, reduce administrative burden, and improve responsiveness, particularly for smaller or ad-hoc purchases. Some viewed direct payments as a way to enhance personalisation and give individuals greater control over their care.<br>*Eligibility*<br>Concerns were raised about offering direct payments to vulnerable groups such as people with substance misuse issues. Risks related to relapse, exploitation, and financial management were cited as reasons for caution. Suggestions for safeguards included using care navigators or trusted third parties to manage funds, or requiring abstinence before eligibility.<br>At the same time, some felt that imposing restrictions such as requiring a sustained period of abstinence from drugs or alcohol, or insisting that personal health budgets be managed by a third party rather than the individual, risked being discriminatory.<br>In contrast, others viewed direct payments as a vital tool for promoting autonomy and personal responsibility. They argued that, with appropriate safeguards in place, individuals should be supported to manage their own budgets wherever possible. |
| Glendinning et al. 2013<br>Personal Health Budgets and Maternity Care: A Qualitative Evaluation. DH 2541. PSSRU, Personal Social Services Research Unit, University of Kent.<br>https://www.phbe.org.uk/documents/DH2541_maternity_report.pdf | Explores the use of PHBs in maternity care through in-depth case studies and interviews | Pregnant women and new mothers from one PHB pilot site focused on maternity care | No details on direct payments in this report. |

Key: PHB: personal health budgets

**Table 4:** Summary of personal health budgets evaluation (National rollout)

| Citation | Content Summary | Population | Direct Payment Findings |
| --- | --- | --- | --- |
| Jones et al. 2017.<br>Personal health budgets: Process and context following the national pilot programme. Working paper 2947. Personal Social Services Research Unit, University of Kent.<br>https://www.pssru.ac.uk/publications/pub-5331/ | The report explores how PHBs have been used in England post-pilot, focusing on their effectiveness, influencing factors, and stakeholder perceptions | Twenty-three PHB users who received their budget following the national pilot and evaluation<br>Eight organisational representatives whose work involved the delivery of personal health budgets within clinical commissioning groups | Over half of the 23 interviewed budget holders managed their PHBs via direct payments.<br>*Personalisation*<br>Users valued the **control** and improved health outcomes direct payments offered.<br>*Factors that enable or hinder successful implementation*<br>Managing direct payments was stressful, especially for those without prior managerial experience. It involved significant paperwork and legal responsibilities. Best suited for individuals who are organised, assertive, and have prior experience in managing responsibilities. Budget holders emphasised the importance of professional support in managing their budgets. Pilot participants noted reduced support after the programme ended, while new budget holders reported little to no support.<br>*Conclusion on PHB in general*<br>Many personal health budget holders highlighted the importance of having support and guidance from professionals around how and on what they could use their budget.<br>There were variations in how budgets could be used, reflecting different interpretations of what counts as meeting needs and improving outcomes.<br>Some budget holders faced restrictions on spending, such as on alternative therapies. Interviews suggested that rules had become stricter over time.<br>Balancing flexibility with guidance remains a challenge, with suggestions including allowing some trial and error and recognising broader definitions of wellbeing.<br>Personal health budgets offered potential empowerment, but interviews suggested this depended on the budget holder's commitment, capacity, and willingness. |
|  |  |  | The effort involved in managing services and planning care was seen as challenging, making the approach more suitable for those able to take on these responsibilities. |
| Jones et al. 2018.<br>Personal health budgets: targeting of support and the service provider landscape. Working Paper 2948. Personal Social Services Research Unit, University of Kent.<br>https://www.pssru.ac.uk/publications/pub-5434/ | The report evaluates the implementation of PHBs in England following the national pilot, with a focus on service provider perspectives, budget holder experiences, and the evolving policy context | Adults supported through NHS CHC, mental health services, learning disability services, and end-of-life care<br>The population reflected broader eligibility criteria compared to the original pilot phase, aligning with the expansion described in the NHS Five Year Forward View and Long Term Plan | The analysis included forty-two PHB support plans. Of these, thirty-one specified the deployment option, with twenty-six indicating that the budget was managed as a direct payment.<br>*Personalisation*<br>**Greater control and flexibility** for budget holders. Enabled **innovative and personalised care**, including non-traditional services like complementary therapies. Improved **independence, dignity, and quality of life** for users and reduced burden on informal carers.<br>*Factors that enable or hinder successful implementation*<br>Managing direct payments was burdensome, especially without prior experience or adequate support. Professional guidance had declined since the pilot, making it harder for users to manage budgets effectively. Local variation in what PHBs could fund led to confusion and inequity. Limited awareness among patients and providers hindered service development and uptake. |

Key: CHC: continuing health care, PHB: personal health budgets

**Table 5:** Summary of organisational reports that explore direct payments.

| Citation | Content Summary | Population | Direct Payment Findings |
| --- | --- | --- | --- |
| The Health Foundation 2011<br>Improvement in practice: the personal touch: the Dutch experience of personal health budgets.<br>https://www.health.org.uk/sites/default/files/ThePersonalTouchDutchExperienceOfPersonalHealthBudgets.pdf | Case study report. It includes interviews with policymakers, carers, users, and experts, and is intended to inform UK policy development | It explores the implementation of PHBs in the Netherlands, aiming to supplement formal evidence with real-world insights<br>At the time of publication PHBs were being piloted in England. This case study report was written to inform policy makers using the case study in the Netherlands as an example | Direct payments are a core feature of the Dutch PHB system. Funds are transferred directly to individuals, who then choose and pay for their own care providers<br>*Implementation Challenges*<br>• This model offers freedom and personalisation, but also complexity—users must manage employment law, budgeting, and quality assurance<br>• The system has led to the rise of intermediary agencies, some of which have been problematic (e.g., fraud, aggressive marketing) and as a result the Dutch government has banned direct payments to intermediaries and introduced a voluntary code of practice |
| Hatton and Waters 2015<br>Personal Health budget holders and family carers. The POET surveys 2015<br>http://s557941885.web.sitehome.co.uk/wp-content/uploads/2018/01/Personal-Health-Budget-Holders-and-Family-Carers-2015-Report.pdf | Evaluation Report. Survey conducted using the Personal Outcomes Evaluation Tool (POET) tool developed by In Control and Lancaster University, and commissioned by NHS England and Think Local Act Personal. | This report evaluates the experiences and outcomes of 302 personal health budget holders and 247 family carers across 31 NHS and council organisations in 37 areas of England<br>The report exclusively examines personal health budgets, which were delivered through direct payments, notional budgets, and third-party arrangements. | *Use of personal budgets*<br>• People most commonly managed their personal health budget through direct payments paid to them (36.7%), followed by direct payments looked after by a friend or family member (26.2%).<br>• Personal health budgets managed by a service provider (13.1%), council or NHS-managed personal health budgets (11.1%) and personal health budgets managed by a broker (11.5%) were less common.<br>*Factors that enable or hinder successful implementation*<br>• People who received a direct payment paid directly to themselves were more likely to plan on their own without any help ($p=0.038$).<br>• People whose direct payment was managed by a family member or friend were more likely to receive planning support from family or friends ($p < 0.001$).<br>• People whose direct payment was managed by a broker were more likely to receive planning support from the council ($p=0.005$).<br>• People who received a direct payment paid directly to themselves were more likely to say it was easy to manage their support ($p=0.049$) and to get the support they wanted ($p=0.004$).<br>• People whose direct payment was managed by a broker were more likely to say it was easy to change their support ($p=0.028$). |
|  |  |  | • People with a direct payment paid directly to themselves were less likely to report a positive impact of their budget on where and with whom they lived, and on feeling safe.<br>• People with a direct payment paid to a family member or friend were more than twice as likely to report that their budget had a positive impact on where and with whom they lived, and on being supported with dignity |
| Skills for Care 2016<br>Support for personal health budget holders who employ personal assistants<br>https://www.skillsforcare.org.uk/Documents/PHB/PHB-Report-Final-2016.pdf<br>Skills for Care 2016<br>Support for personal health budget holders who employ personal assistants: Executive summary<br>https://www.skillsforcare.org.uk/Documents/PHB/PHB-Report-Exec-Summary-2016.pdf | Research and case study-based evaluation exploring how personal health budgets using direct payments to employ personal assistants are supported across health and care systems. | This report evaluates the experiences of personal health budget holders who manage their budget via direct payments to employ personal assistants. It draws on 60 survey responses covering approximately 50 clinical commissioning group areas in England, as well as interviews with individuals and teams from 10 participating sites. Detailed evidence from seven of these sites was used for in-depth case study analysis | Recommendations from executive summary<br>*Training*<br>• Clinical commissioning groups should develop local frameworks for delegating healthcare tasks, including training and competence assessment. Clinical commissioning groups should offer core training for personal assistants, linked where appropriate to the Care Certificate standards.<br>• Clinical commissioning groups should plan for increased demand by creating roles to oversee training, sign-off, and review of competence<br>*Support for Personal Assistants*<br>• Personal assistants should have access to peer support or, where not feasible, a neutral point of contact for workplace or human resources issues, separate from employer support<br>*Recruitment and Retention*<br>• National Health Service organisations, local authorities, and other stakeholders should work together to address recruitment and retention challenges<br>• National work is needed to understand how new personal assistant employment models may affect current systems |
| Alakeson and Rumbold 2013. Personal health budgets: Challenges for commissioners and policy-makers. London: The Nuffield Trust.<br>https://www.nuffieldtrust.org.uk/research/personal-health-budgets-challenges-for-commissioners-and-policy-makers | Research summary using the evidence from the national pilot programme launched in 2009 to test the feasibility and impact of PHB's. | The national evaluation was conducted across 20 sites covering various long-term conditions including mental health, stroke, and NHS continuing healthcare. The report was intended for commissioners and policy-makers in the UK health system to inform a wider roll-out of PHBs by highlighting practical, financial, and policy challenges | Direct Payments:<br>• Direct payment support services are available to help people with the responsibilities of being an employer such as payroll facilities or third-party arrangements can be used for those who cannot or do not want to manage the financial responsibility directly<br>Key points from the research summary:<br>Implementing is the responsibility of clinical commissioning groups: the summary provides important issues for commissioners:<br>*Decommissioning existing services*<br>• Commissioners must plan to phase out services not chosen by PHB holders to avoid duplication and market shrinkage.<br>*Developing and diversifying the provider market*<br>• Efforts are needed to diversify and grow the market of providers needed whilst avoiding destabilising existing providers.<br>• There are a range of transition strategies that commissioners can use to support the provider market.<br>*Funding the necessary infrastructure*<br>• New infrastructure around budget setting, care planning and system monitoring is required with some evidence to suggest that some efficiency can be achieved by 'piggy-backing' systems that already exist<br>• Bringing PHBs together with personal budgets in social care to create integrated individual budgets potentially offers a new route to service integration. The research summary refers to a 'dual carriageway' approach which brings together the referral, assessment, budget setting, planning and monitoring of different budgets without the complexities of structural integration between organisations and government departments<br>Policy-makers need to be aware that there is a risk of a postcode lottery emerging |

Key: PHB: Personal Health budgets

## 7.2 Acknowledgments

The authors would like to thank Nia Griffiths, Lisa Bridges, Emily Keoghane and Beti-Jane Ingram for their contributions during stakeholder meetings in guiding the focus of the review and interpretation of findings.

## 8. APPENDIX 1: Resources searched during Rapid Evidence Summary

A single list of resources has been developed for guiding and documenting the sources searched as part of Rapid Evidence Summary.

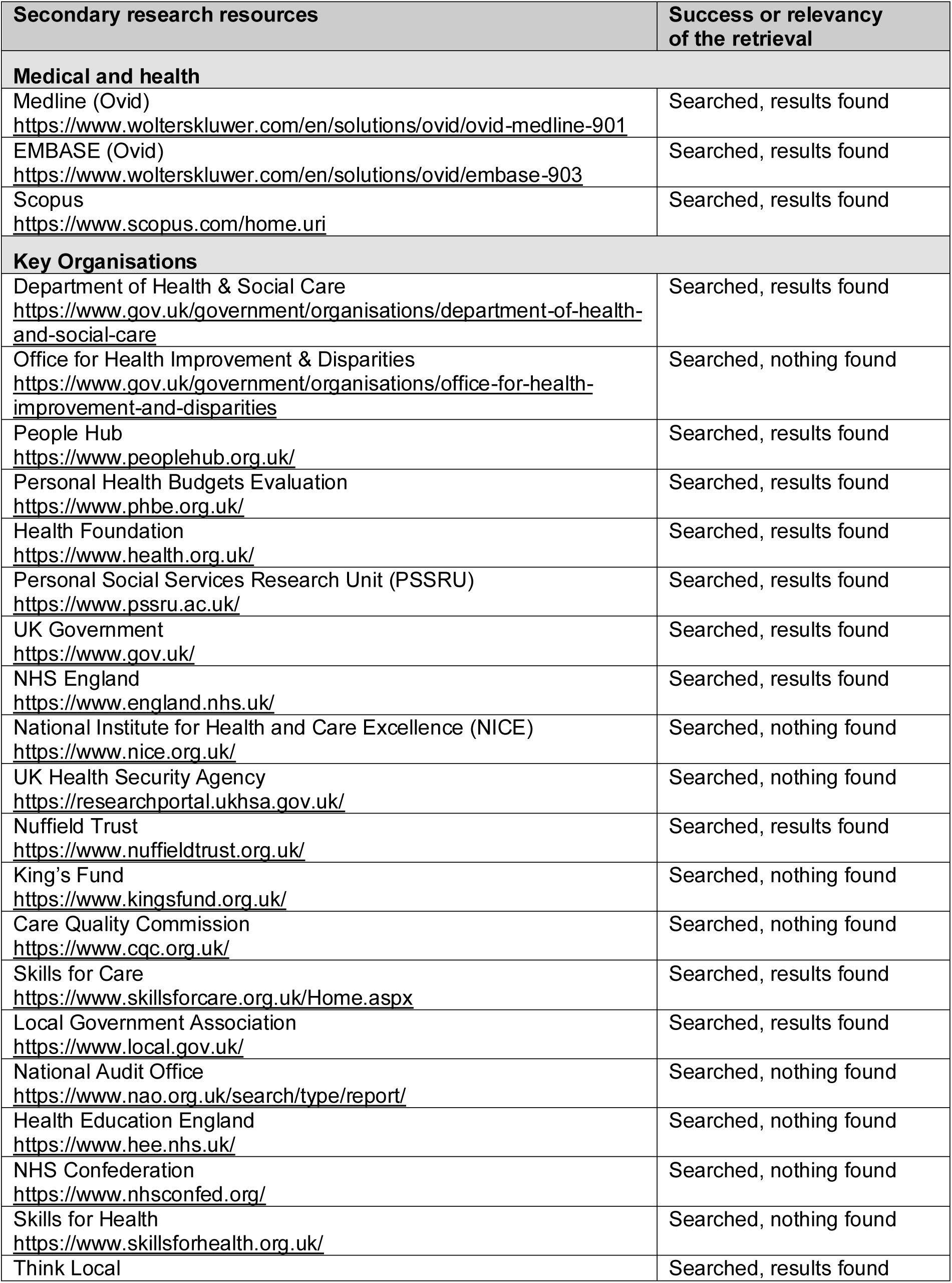
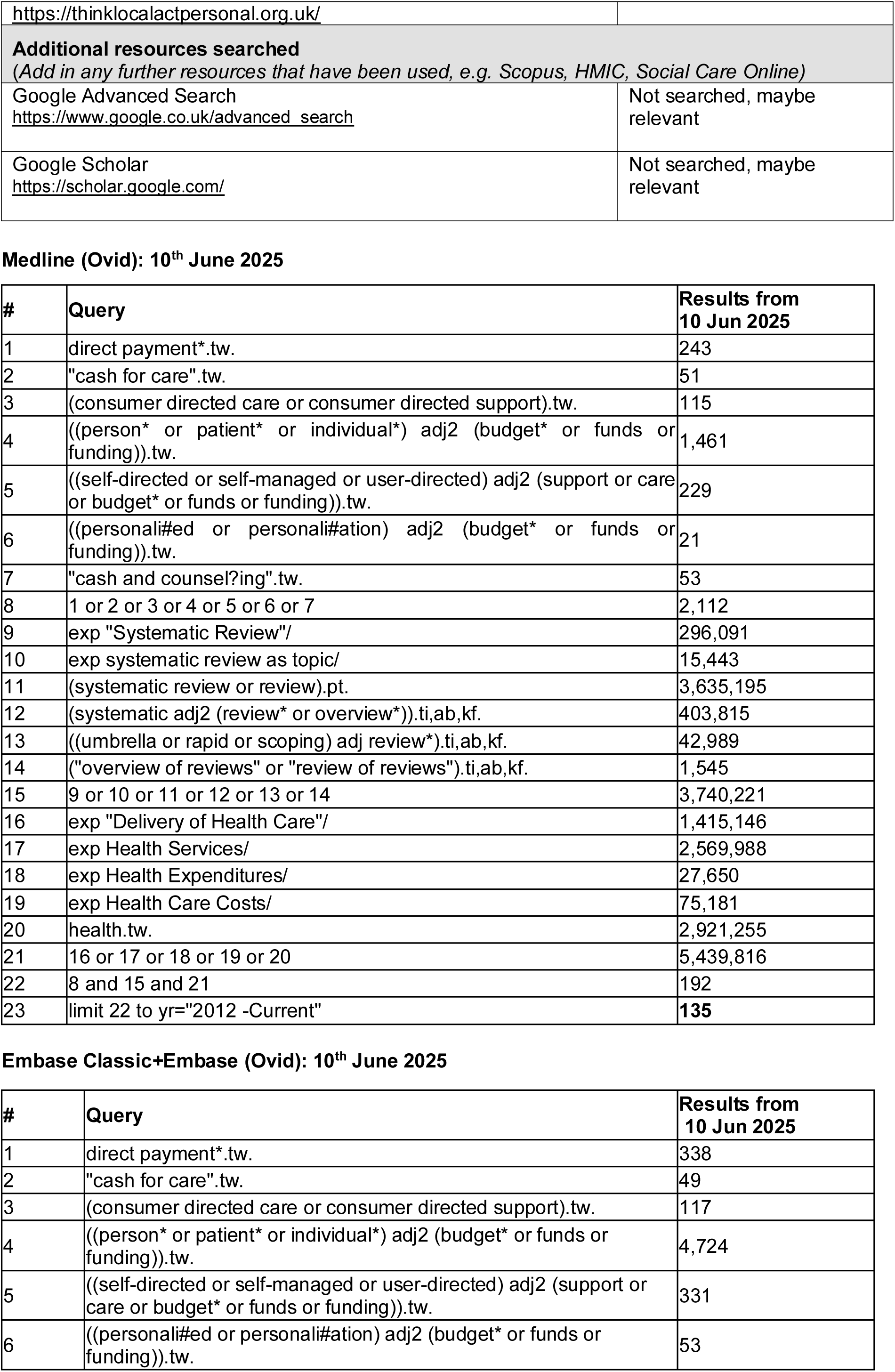
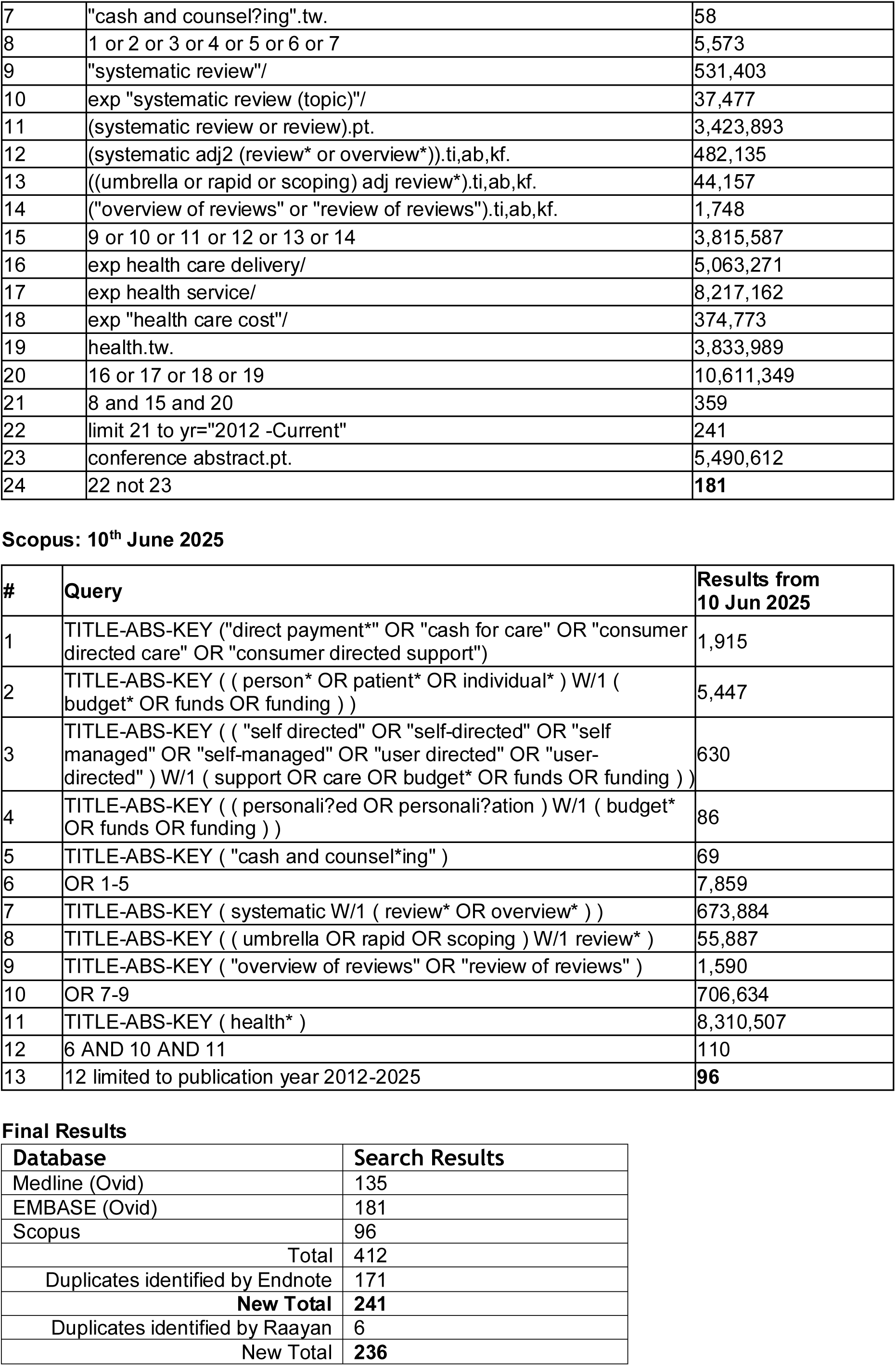

## Footnotes

2 *Open model -* Payment for care is provided to those eligible for long-term care services with few strings attached. The cash allowance can be spent however the recipient chooses and the money does not have to be accounted for (Alakeson 2010) *Budgeted or planned models -* Programme maintains a more direct connection between a participant’s needs and the goods and services purchased to meet those needs. Restrictions are placed on how money can be spent, and the expenditure is audited carefully (Alakeson 2010)

3 Carers and personal assistants – friends relatives or previous care staff

